# Neutralising antibodies in Spike mediated SARS-CoV-2 adaptation

**DOI:** 10.1101/2020.12.05.20241927

**Authors:** SA Kemp, DA Collier, R Datir, IATM Ferreira, S Gayed, A Jahun, M Hosmillo, C Rees-Spear, P Mlcochova, Ines Ushiro Lumb, David J Roberts, Anita Chandra, N Temperton, The CITIID-NIHR BioResource COVID-19 Collaboration, The COVID-19 Genomics UK (COG-UK) Consortium, K Sharrocks, E Blane, JAG Briggs, MJ van Gils, KGC Smith, JR Bradley, C Smith, R Doffinger, L Ceron-Gutierrez, G Barcenas-Morales, DD Pollock, RA Goldstein, A Smielewska, JP Skittrall, T Gouliouris, IG Goodfellow, E Gkrania-Klotsas, CJR Illingworth, LE McCoy, RK Gupta

**Affiliations:** Division of Infection and Immunity, University College London, London, UK; Cambridge Institute of Therapeutic Immunology & Infectious Disease (CITIID), Cambridge, UK; Department of Medicine, University of Cambridge, Cambridge, UK; Department of Infectious Diseases, Cambridge University NHS Hospitals Foundation Trust, Cambridge, UK; Department of Pathology, University of Cambridge, Cambridge; NHS Blood and Transplant, Oxford and BRC Haematology Theme, University of Oxford, UK; Viral Pseudotype Unit, Medway School of Pharmacy, University of Kent, UK; Medical Research Council Laboratory of Molecular Biology, Cambridge, UK; Department of Medical Microbiology, Academic Medical Center, University of Amsterdam, Amsterdam Institute for Infection and Immunity, Amsterdam, Netherlands; NIHR Cambridge Clinical Research Facility, Cambridge, UK; Department of Virology, Cambridge University NHS Hospitals Foundation Trust; Department of Applied Mathematics and Theoretical Physics, University of Cambridge, UK; Department of Clinical Biochemistry and Immunology, Addenbrookes Hospital; Biochemistry and Molecular Genetics, University of Colorado School of Medicine, Aurora, Colorado, USA; Clinical Microbiology and Public Health Laboratory, Addenbrookes’ Hospital, Cambridge, UK; MRC Biostatistics Unit, University of Cambridge, Cambridge, UK; Africa Health Research Institute, Durban, South Africa

**Keywords:** SARS-CoV-2, COVID-19, antibody escape, Convalescent plasma, neutralising antibodies, mutation, evasion, resistance, immune suppression

## Abstract

SARS-CoV-2 Spike protein is critical for virus infection via engagement of ACE2, and amino acid variation in Spike is increasingly appreciated. Given both vaccines and therapeutics are designed around Wuhan-1 Spike, this raises the theoretical possibility of virus escape, particularly in immunocompromised individuals where prolonged viral replication occurs. Here we report chronic SARS-CoV-2 with reduced sensitivity to neutralising antibodies in an immune suppressed individual treated with convalescent plasma, generating whole genome ultradeep sequences by both short and long read technologies over 23 time points spanning 101 days. Although little change was observed in the overall viral population structure following two courses of remdesivir over the first 57 days, N501Y in Spike was transiently detected at day 55 and V157L in RdRp emerged. However, following convalescent plasma we observed large, dynamic virus population shifts, with the emergence of a dominant viral strain bearing D796H in S2 and **Δ**H69/**Δ**V70 in the S1 N-terminal domain NTD of the Spike protein. As passively transferred serum antibodies diminished, viruses with the escape genotype diminished in frequency, before returning during a final, unsuccessful course of convalescent plasma. In vitro, the Spike escape double mutant bearing **Δ**H69/**Δ**V70 and D796H conferred decreased sensitivity to convalescent plasma, whilst maintaining infectivity similar to wild type. D796H appeared to be the main contributor to decreased susceptibility, but incurred an infectivity defect. The **Δ**H69/**Δ**V70 single mutant had two-fold higher infectivity compared to wild type and appeared to compensate for the reduced infectivity of D796H. Consistent with the observed mutations being outside the RBD, monoclonal antibodies targeting the RBD were not impacted by either or both mutations, but a non RBD binding monoclonal antibody was less potent against **Δ**H69/**Δ**V70 and the double mutant. These data reveal strong selection on SARS-CoV-2 during convalescent plasma therapy associated with emergence of viral variants with reduced susceptibility to neutralising antibodies.

## Introduction

SARS-CoV-2 is an RNA betacoronavirus, with closely related viruses identified in pangolins and bats^1,2^. RNA viruses have inherently higher rates of mutation than DNA viruses such as Herpesviridae^3^. However, amongst RNA viruses coronaviruses have a relatively modest mutation rate at around 23 nucleotide substitutions per year^4^, likely due to proof reading capability of coronavirus RNA dependent RNA polymerase^5^. The capacity for successful adaptation is exemplified by the Spike D614G mutation, that arose in China and rapidly spread worldwide^6^, now accounting for more than 90% of infections. The mutation appears to increase infectivity and transmissibility in animal models^7^. Although the SARS-CoV-2 Spike protein is critical for virus infection via engagement of ACE2, substantial Spike amino acid variation has been observed in circulating viruses^8^. Mutations in the receptor binding domain (RBD) of Spike are of particular concern because the RBD is an important target of neutralising antibodies and therapeutic monoclonal antibodies.

Deletions in the N-terminal domain (NTD) of Spike S1 have also been increasingly recognised, both within hosts^9^ and across individuals^10^. The evolutionary basis for the emergence of deletions is unclear at present but could be related to escape from immunity or to enhanced fitness/transmission. The most notable deletion in terms of frequency is **Δ**H69/**Δ**V70. This double deletion has been detected in multiple unrelated lineages, including the recent ‘Cluster 5’ mink related strain in the North Jutland region of Denmark (https://files.ssi.dk/Mink-cluster-5-short-report_AFO2). There it was associated with the RBD mutation Y453F in almost 200 individuals. Another European cluster in GISAID includes **Δ**H69/**Δ**V70 along with the RBD mutation N439K.

Although **Δ**H69/**Δ**V70 has been detected multiple times, within-host emergence remains undocumented and the reasons for its selection are unknown. Here we document real time SARS-CoV-2 emergence of **Δ**H69/**Δ**V70 in combination with the S2 mutation D796H following convalescent plasma therapy in an immunocompromised human host, demonstrating selection and reduced phenotypic susceptiblility of selected mutations.

## Results

### Clinical case history of SARS-CoV-2 infection in setting of immune-compromised host

Clinical data are available from the corresponding author.

Given the history of B cell depletion therapy and hypogammaglobulinemia we measured serum SARS-CoV-2 specific antibodies over the course of the admission. Total serum antibodies to SARS-CoV-2 were tested at days 44 and 50 by S protein immunoassay (Siemens). Results were negative. Three units (200mL each) of convalescent plasma (CP) from three independent donors were obtained through the NHS Blood and Transplant Clearance Registry and administered on compassionate named patient basis. These had been assayed for SARS-CoV-2 IgG antibody titres (Supplementary figure 4). Patient serum was subsequently positive for SARS-CoV-2 specific antibodies by S protein immunoassay (Siemens) in the hospital diagnostic laboratory on days 68, 90 and 101.

### Virus genomic comparative analysis of 23 sequential respiratory samples over 101 days

The majority of samples were respiratory samples from nose and throat or endotracheal aspirates during the period of intubation. Ct values ranged from 16-34 and all 23 respiratory samples were successfully sequenced by standard single molecule sequencing approach as per the ARTIC protocol implemented by COG-UK; of these 20 additionally underwent short-read deep sequencing using the Illumina platform (Supplementary Table 3). There was generally good agreement between the methods (Supplementary Figure 5, Supplementary Table 4). Additionally, Single genome amplification and sequencing of Spike using extracted RNA from respiratory samples was used as an independent method to detect mutations observed (Supplementary Table 5). Finally, we detected no evidence of recombination, based on two independent methods.

Maximum likelihood analysis of patient-derived whole genome consensus sequences demonstrated clustering with other local sequences from the same region (Figure 2A). The infecting strain was assigned to lineage 20B bearing the D614G Spike variant. Environmental sampling showed evidence of virus on surfaces such as telephone and call bell. Sequencing of these surface viruses showed clustering with those derived from the respiratory tract (Figure 2B). All samples were consistent with having arisen from a single viral population. In our phylogenetic analysis, we included sequential sequences from three other local patients identified with persistent viral RNA shedding over a period of 4 weeks or more as well as two long term immunosuppressed SARS-CoV-2 ‘shedders’ recently reported^9,11^, (Figure 2B and Supplementary Table 2). While the sequences from the three local patients as well as from Avanzato et al^11^ showed little divergence with no amino acid changes in Spike over time, the case patient showed significant diversification. The Choi^9^ et al report showed similar degree of diversification as the case patient. Further investigation of the sequence data suggested the existence of an underlying structure to the viral population in our patient, with samples collected at days 93 and 95 being rooted within, but significantly divergent from the original population (Figure 2B and 3). The relationship of the divergent samples to those at earlier time points argues against superinfection.

**Figure 1:**
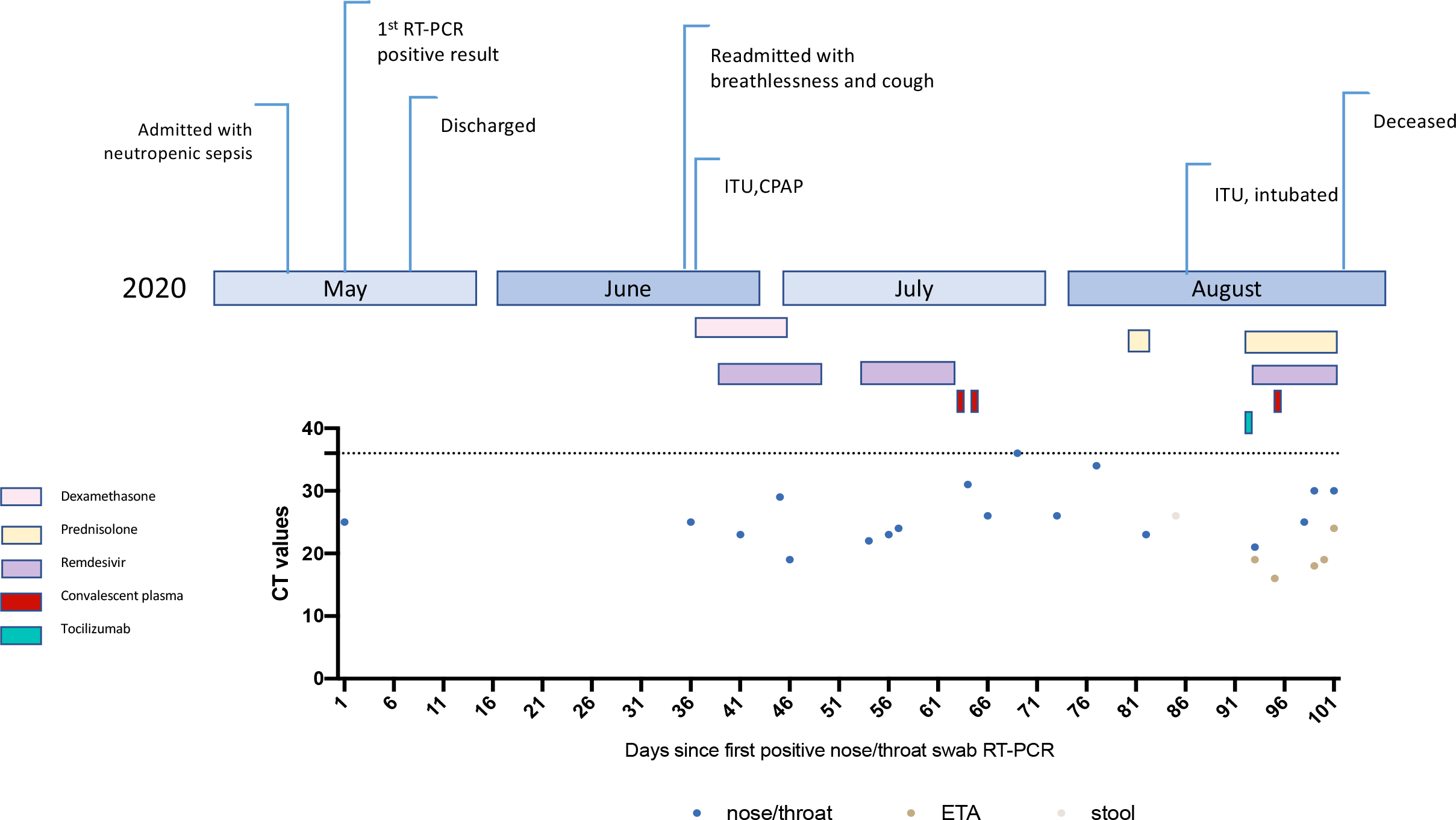
Clinical time line of events with longitudinal respiratory sample CT values. CT-cycle threshold.

**Figure 2.**
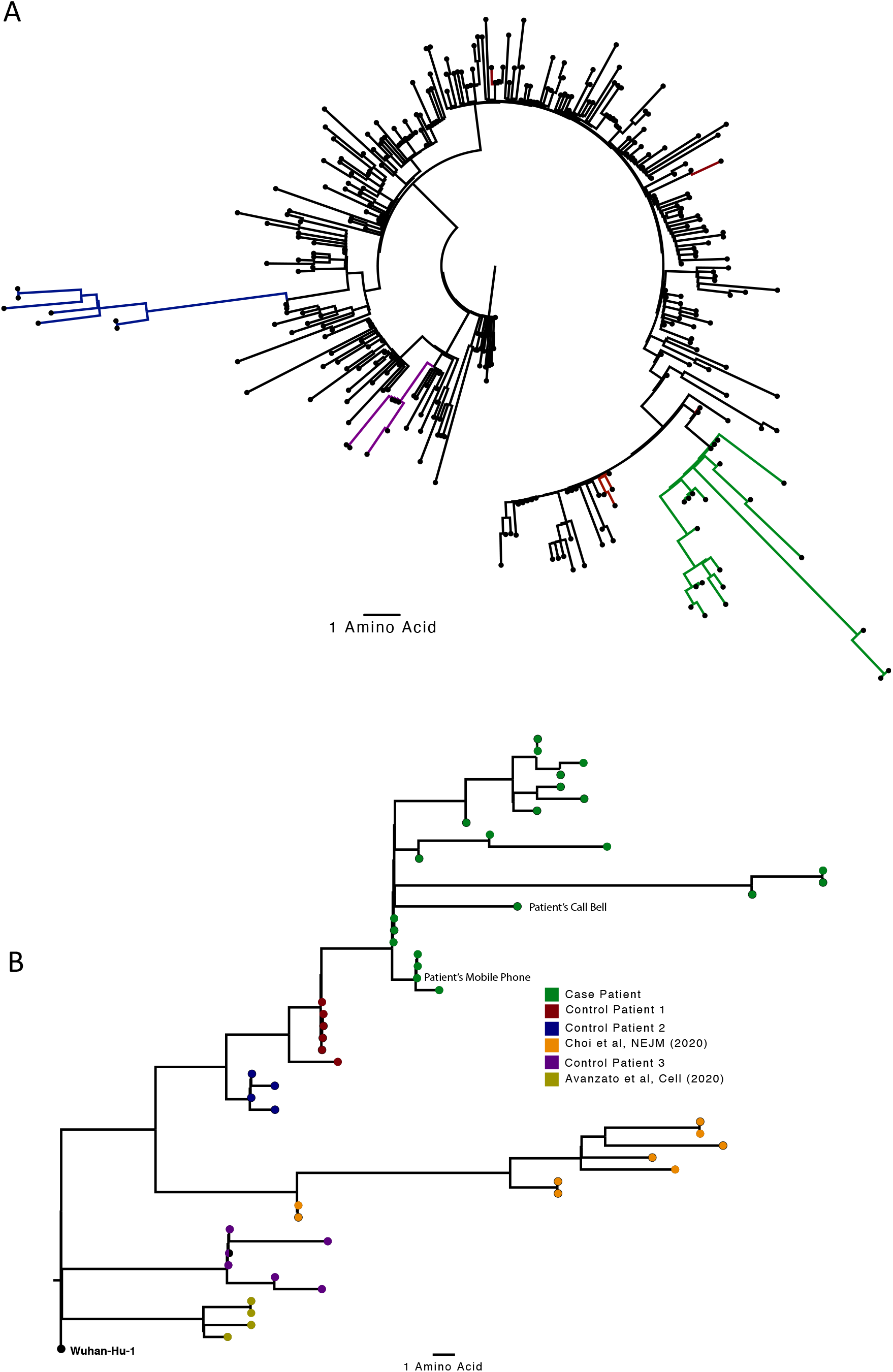
Analysis of 23 Patient derived whole SARS-CoV-2 genome sequences in context of national sequences and other cases of chronic SARS-CoV-2 shedding. A. Circularised maximum-likelihood phylogenetic tree rooted on the Wuhan-Hu-1 reference sequence, showing a subset of 250 local SARS-CoV-2 genomes from GISAID. This diagram highlights significant diversity of the case patient (green) compared to three other local patients with prolonged shedding (blue, red and purple sequences). All “United Kingdom / English” SARS-CoV-2 genomes were downloaded from the GISAID database and a random subset of 250 selected as background. **B**. Close-view maximum-likelihood phylogenetic tree indicating the diversity of the case patient and three other long-term shedders from the local area (red, blue and purple), compared to recently published sequences from Choi et al (orange) and Avanzato et al (gold). Control patients generally showed limited diversity temporally, though the Choi et al sequences were found to be even more divergent than the case patient. Environmental samples (patient’s call bell, and patient’s mobile phone) are indicated.

### SARS-CoV-2 antibodies in serum, and convalescent plasma associated changes in viral diversity

We measured longitudinal serum and convalescent plasma SARS-CoV-2 IgG to SARS-CoV-2 trimeric Spike (S), Spike receptor binding domain (RBD) and Nucleocapsid protein (N) using a Luminex based assay. Levels of antibodies were very low at day 39, consistent with the standard lab based assay result above. Following CP1 at day 63 and CP2 at day 65, antibody levels were significantly increased against all three protein targets. As expected antibody titres fell over time before increasing after the third unit of CP on day 93.

All samples tested positive by RT-PCR and there was no sustained change in Ct values throughout the 101 days following the first two courses of remdesivir (days 41 and 54), or the first two units of convalescent plasma (days 63 and 65). Consensus sequences from short read deep sequence Illumina data revealed dynamic population changes after day 65, as shown by a highlighter plot (Figure 3). In addition, we were also able to follow the dynamics of virus populations down to low frequencies during the entire period (Figure 4). Following remdesivir at day 41 the low frequency variant analysis allowed us to observe transient amino acid changes in populations at below 50% abundance in Orf 1b, 3a and Spike, with aT39I mutation in ORF7a reaching 77% on day 45 (Figure 4). At day 66 we noted I513T in NSP2 and V157L in RdRp had emerged from undetectable at day 54 to 100% frequency (Figure 4 orange line), with the polymerase being the more plausible candidate for driving this sweep. Notably, spike variant N501Y, which can increase the ACE2 receptor affinity^12^, and which is present in the new UK B1.1.7 lineage^13^, was observed on day 55 at 33% frequency, but was eliminated by the sweep of the NSP2/RdRp variant.

**Figure 3.**
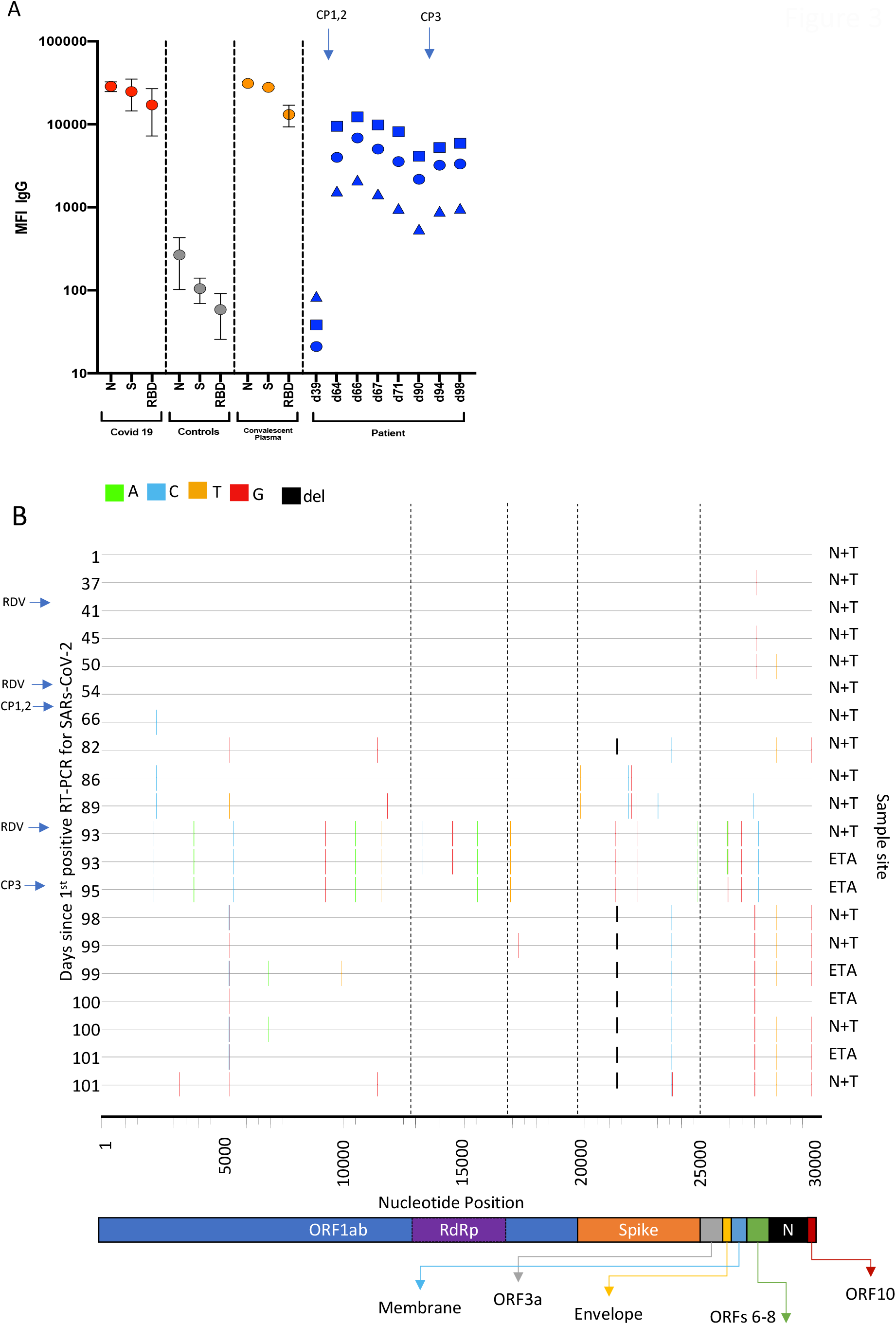
Serum SARS-CoV-2 antibody levels and virus population changes in chronic SARS-CoV-2 infection. A. Anti SARS-CoV2 IgG antibodies in patient and pre/post convalescent plasma compared to RNA+ Covid19 patients and prepandemic healthy controls: Red, grey and gold: IgG antibodies to SARS-CoV2 nucleocapsid protein (N), trimeric S protein (S) and the receptor binding domain (RBD) were measured by multiplexed particle based flow cytometry (Luminex) in RNA+ Covid 19 patients (N=20, red dots), Pre-pandemic healthy controls (N=20, grey dots) and in the convalescent donor plasma (orange dots); Results are shown as mean fluorescent intensity (MFI) +/- SD. **Patient sera over time in blue:** Anti SARS-CoV2 IgG to N (blue squares), S (blue circles) and RBD (blue triangles). Timing of CP units is also shown **B. Highlighter plot indicating nucleotide changes at consensus level in sequential respiratory samples compared to the consensus sequence at first diagnosis of COVID-19**. Each row indicates the timepoint the sample was collected (number of days from first positive SARS-CoV-2 RT-PCR). Black dashed lines indicate the RNA-dependent RNA polymerase (RdRp) and Spike regions of the genome. There were few nucleotide substitutions between days 1-54, despite the patient receiving two courses of remdesivir. The first major changes in the spike genome occurred on day 82, following convalescent plasma given on days 63 and 65. The amino acid deletion in S1, ΔH69/V70 is indicated by the black lines. Sites: Endotracheal aspirate (ETA) or Nose/throat swabs (N+T).

**Figure 4.**
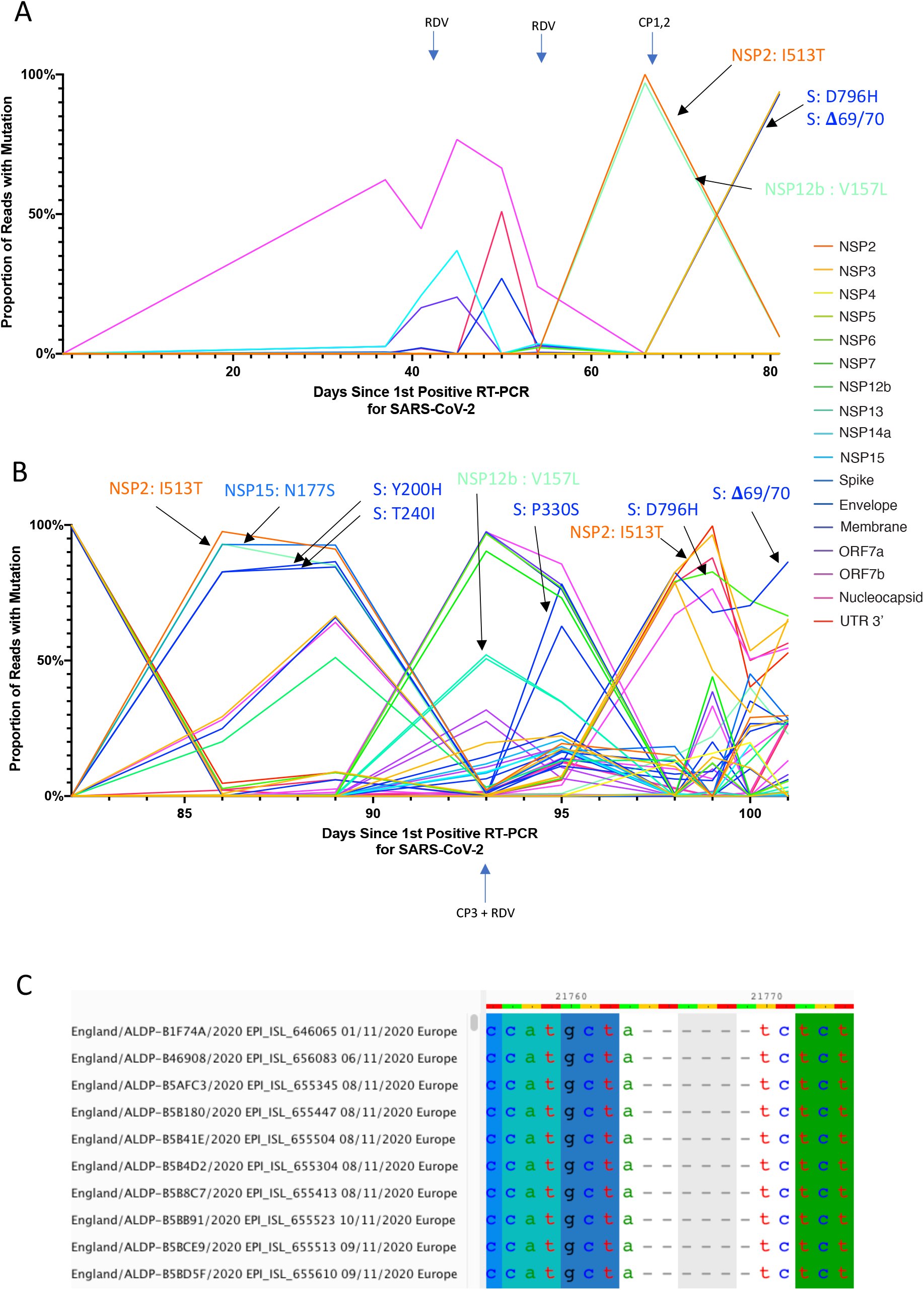
Whole genome variant trajectories showing amino acids and relationship to treatments. Data based on Illumina short read ultra deep sequencing at 1000x coverage. Variants shown reached a frequency of at least 10% in at least 2 samples. Treatments indicated are convalescent plasma (CP) and Remdesivir (RDV). Variants described in the text are designated by labels using the same colouring as the position in the genome. **A**. Variants detected in the patient from days 1-81. **B**. Variants detected in the patient from day 81-101. **C**. Multiple sequence alignment of 10 randomly selected SARS-CoV-2 Genomes containing the **Δ**69/70 deletion. In all cases, six nucleotides are deleted that are out of frame.

In contrast to the early period of infection, between days 66 and 82, following the first two administrations of convalescent sera, a dramatic shift in the virus population was observed, with a variant bearing D796H in S2 and **Δ**H69/**Δ**V70 in the S1 N-terminal domain (NTD) becoming the dominant population at day 82. This was identified in a nose and throat swab sample with high viral load as indicated by Ct of 23 (Figure 5). The deletion was not detected at any point prior to the day 82 sample, even as minority variants by short read deep sequencing.

**Figure 5.**
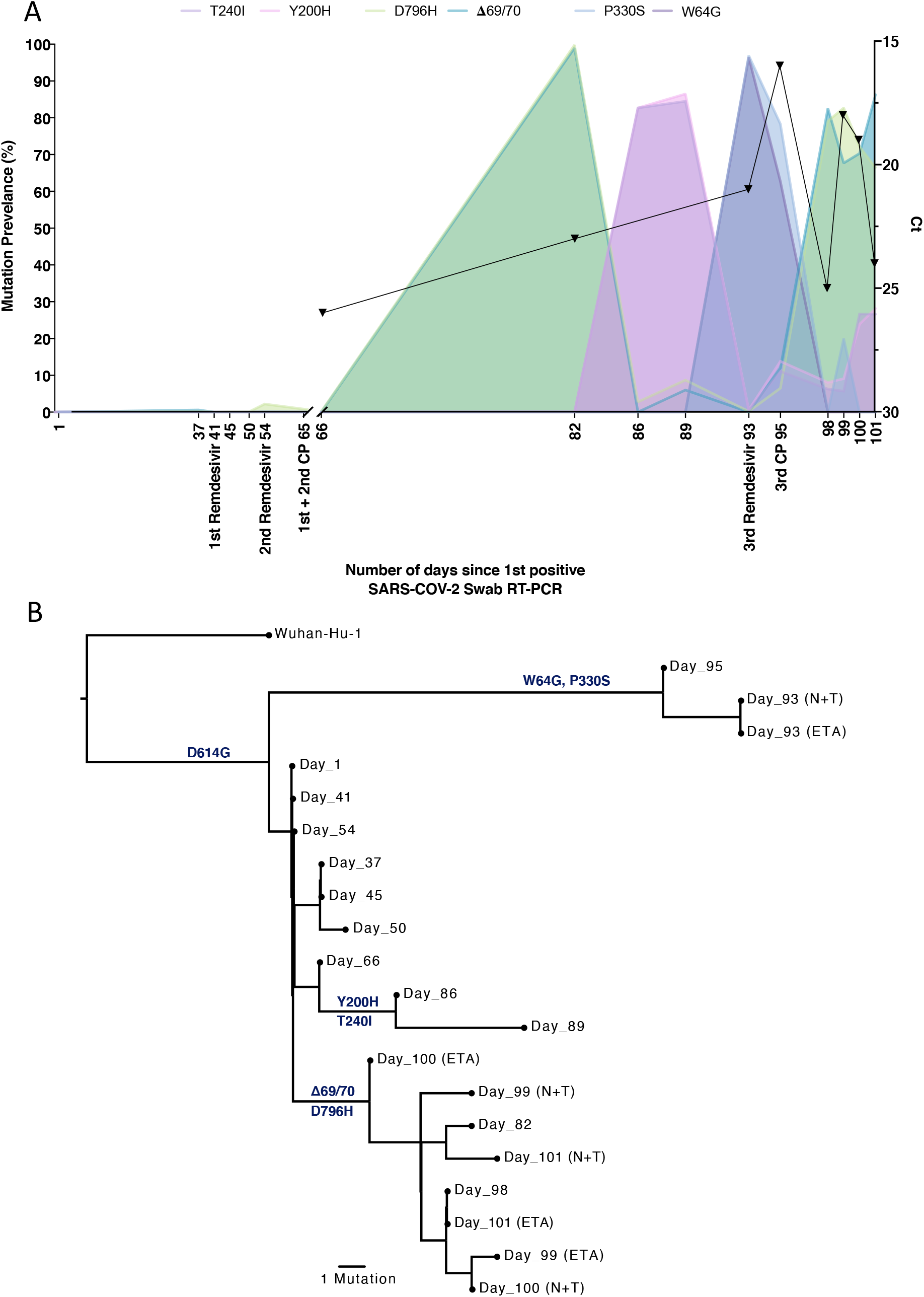
Longitudinal variant frequencies and phylogenetic relationships for virus populations bearing six Spike (S) mutations A. At baseline, all four S variants (Illumina sequencing) were absent (<1% and <20 reads). Approximately two weeks after receiving two units of convalescent plasma (CP), viral populations carrying ΔH69/V70 and D796H mutants rose to frequencies >90% but decreased significantly four days later. This population was replaced by a population bearing Y200H and T240I, detected in two samples over a period of 6 days. These viral populations were then replaced by virus carrying W64G and P330S mutations in Spike, which both dominated at day 93. Following a 3rd course of remdesivir and an additional unit of convalescent plasma, the ΔH69/V70 and D796H virus population re-emerged to become the dominant viral strain reaching variant frequencies of >90%. Pairs of mutations arose and disappeared simultaneously indicating linkage on the same viral haplotype. CT values from respiratory samples are indicated on the right y-axis (black triangles) **B**. Maximum likelihood phylogenetic tree of the case patient with day of sampling indicated. Spike mutations defining each of the clades are shown ancestrally on the branches on which they arose. On dates where multiple samples were collect, these are indicated as endotracheal aspirate (ETA) and Nose + throat swabs (N+T).

On Days 86 and 89, viruses obtained from upper respiratory tract samples were characterised by the Spike mutations Y200H and T240I, with the deletion/mutation pair observed on day 82 having fallen to frequencies of 10% or less (Figure 4 and 5). The Spike mutations Y200H and T240I were accompanied at high frequency by two other non-synonymous variants with similar allele frequencies, coding for I513T in NSP2, V157L in RdRp and N177S in NSP15 (Figure 4). Of these, the former was previously observed at 100% frequency in the sample on day 66 (Figure 3, orange line), arguing that this new lineage emerged out of a previously-existing population.

Sequencing of a nose and throat swab sample at day 93 identified viruses characterised by Spike mutations P330S at the edge of the RBD and W64G in S1 NTD at close to 100% abundance, with D796H along with **Δ**H69/**Δ**V70 at <1% abundance and the variants Y200H and T240I at frequencies of <2%. Viruses with the P330S variant were detected in two independent samples from different sampling sites, arguing against the possibility of contamination. The divergence of these samples from the remainder of the population (Figure 4, 5B and Supplementary Figure 6) suggests the possibility that they represent the emergence of a previously unobserved subpopulation. Following the third course of remdesivir (day 93) and third CP (day 95), we observed a re-emergence of the D796H + **Δ**H69/**Δ**V70 viral population. The inferred linkage of D796H and **Δ**H69/**Δ**V70 was maintained as evidenced by the highly similar frequencies of the two variants.

Patterns in the variant frequencies suggest competition between virus populations carrying different mutations, viruses with the D796H/**Δ**H69/**Δ**V70 deletion/mutation pair rising to high frequency during CP therapy, then being outcompeted by another population in the absence of therapy. Specifically, these data are consistent with a lineage of viruses with the NSP2 I513T and RdRp V157L variant, dominant on day 66, being outcompeted during therapy by the mutation/deletion variant. With the lapse in therapy, the original strain, having acquired NSP15, N1773S and the Spike mutations, regained dominance, followed by the emergence of a separate population with the W64G and P330S mutations.

In a final attempt to reduce the viral load, a third course of remdesivir (day 93) and third CP (day 95) were administered. We observed a re-emergence of the D796H + **Δ**H69/**Δ**V70 viral population. The inferred linkage of D796H and **Δ**H69/**Δ**V70 was maintained as evidenced by the highly similar frequencies of the two variants, suggesting that the third unit of CP led to the re-emergence of this population under renewed positive selection. In further support of our proposed idea of competition, noted above, frequencies of these two variants appeared to mirror changes in the NSP2 I513T mutation (Figure 4), suggesting these as markers of opposing clades in the viral population. Ct values remained low throughout this period with hyperinflammation, eventually leading to multi-organ failure and death at day 102. The repeated increase in frequency of the viral population with CP therapy strongly supports the hypothesis that the deletion/mutation combination conferred selective advantage.

### Spike mutants emerging post convalescent plasma impair neutralisating antibodies

Using lentiviral pseudotyping we generated wild type, **Δ**H69/**Δ**V70 + D796H and single mutant Spike proteins in enveloped virions in order to measure neutralisation activity of CP against these viruses (Figure 6). This system has been shown to give generally similar results to replication competent virus^14,15^. Spike protein from each mutant was detected in pelleted virions (Figure 6A). We also probed with an HIV-1 p24 antibody to monitor levels of lentiviral particle production. We then measured infectivity of the pseudoviruses, correcting for virus input, and found that **Δ**H69/**Δ**V70 appeared to have two-fold higher infectivity over a single round of infection compared to wild type (Figure 6B, supplementary figure 7). By contrast, the D796H single mutant had significantly lower infectivity as compared to wild type and the double mutant had similar infectivity to wild type (Figure 6B, supplementary figure 7).

**Figure 6:**
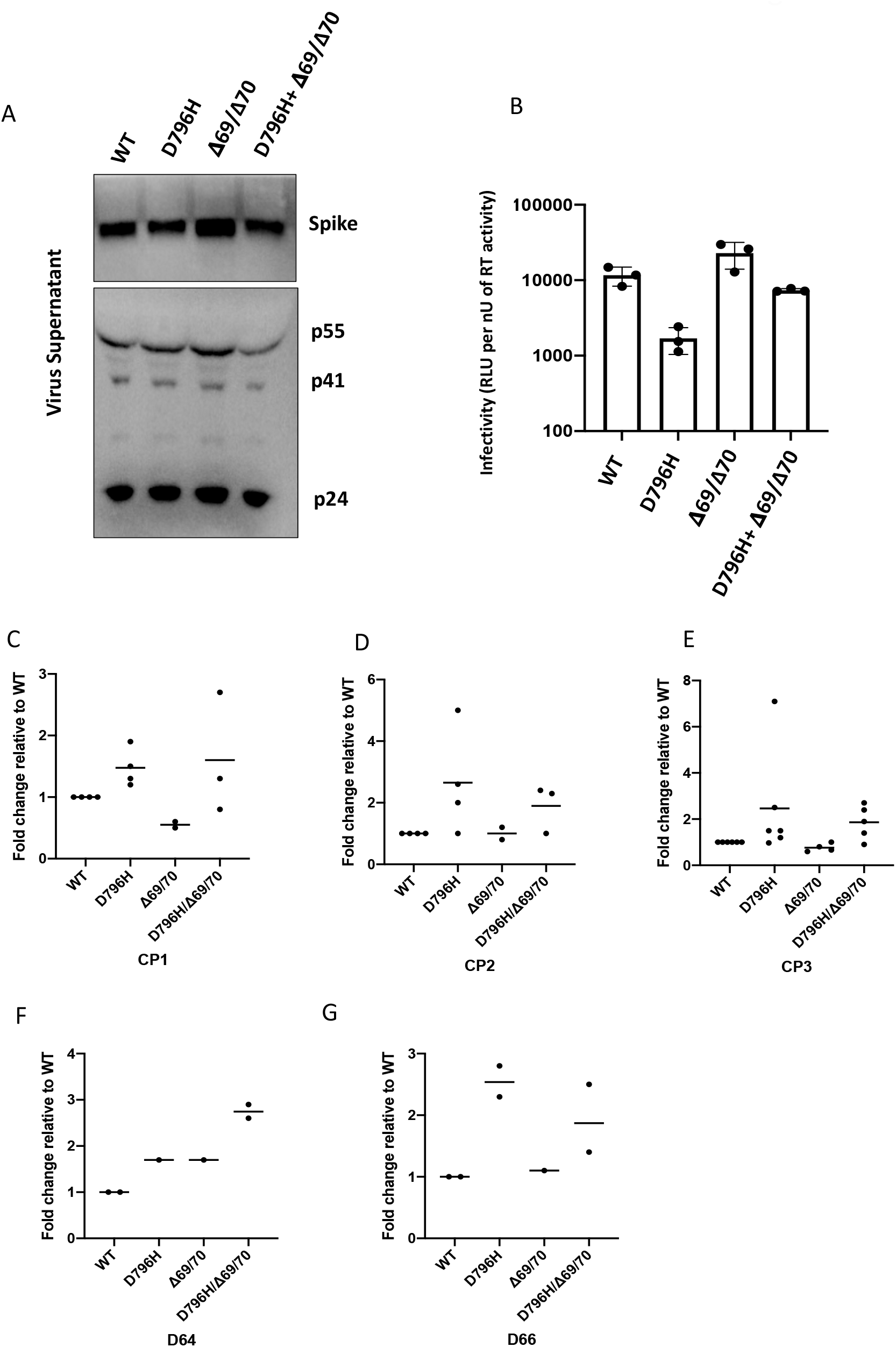
Spike mutant D796H + ΔH69/V70 has infectivity comparable to wild type but is less sensitive to multiple units of convalescent plasma (CP). A. western blot of virus pellets after centrifugation of supernatants from cells transfected with lentiviral pseudotyping plasmids including Spike protein **B**. Single round Infectivity of luciferase expressing lentivirus pseudotyped with SARS-CoV-2 Spike protein (WT versus mutant) on 293T cells co-transfected with ACE2 and TMPRSS2 plasmids. Infectivity is corrected for reverse transcriptase activity in virus supernatant as measured by real time PCR **C-E**. convalescent plasma (CP units 1-3) neutralization potency. against pseudovirus virus bearing Spike mutants D796H, ΔH69/V70 and D796H + ΔH69/V70 F, G patient serum neutralisation potency against pseudovirus virus bearing Spike mutants D796H, ΔH69/V70 and D796H + ΔH69/V70. Patient serum was taken at indicated Day (D). Indicated is serum dilution required to inhibit 50% of virus infection (ID50). Data points represent means of technical replicates and each data point is an independent exp.

We found that D796H alone and the D796H + **Δ**H69/**Δ**V70 double mutant were less sensitive to neutralisation by convalescent plasma samples (Figure 6C-E). By contrast the **Δ**H69/**Δ**V70 single mutant did not impact neutralisation. In addition, patient derived serum from days 64 and 66 (one day either side of CP2 infusion) similarly showed lower potency against the D796H + **Δ**H69/**Δ**V70 mutants (Figure 6F, G).

A panel of nineteen monoclonal antibodies (mAbs) isolated from three donors was previously identified to neutralize SARS-CoV-2. To establish if the mutations incurring in vivo (D796H and **Δ**H69/**Δ**V70) resulted in a global change in neutralization sensitivity we tested neutralising mAbs targeting the seven major epitope clusters previously described (excluding non-neutralising clusters II, V and small [n =<2] neutralising clusters IV, X). The seven RBD-specific mAbs (Supplementary Table 6) exhibited no major change in neutralisation potency and non-RBD specific COVA1-21 showing 3-5 fold reduction in potency against **Δ**H69/**Δ**V70+D796H and **Δ**H69/**Δ**V70, but not D796H alone^15^ (Figure 7). We observed no differences in neutralisation between single/double mutants and wild type, suggesting that the mechanism of escape was likely outside these epitopes in the RBD. These data confirm the specificity of the findings from convalescent plasma and suggest that mutations observed are related to antibodies targeting regions outside the RBD.

**Figure 7:**
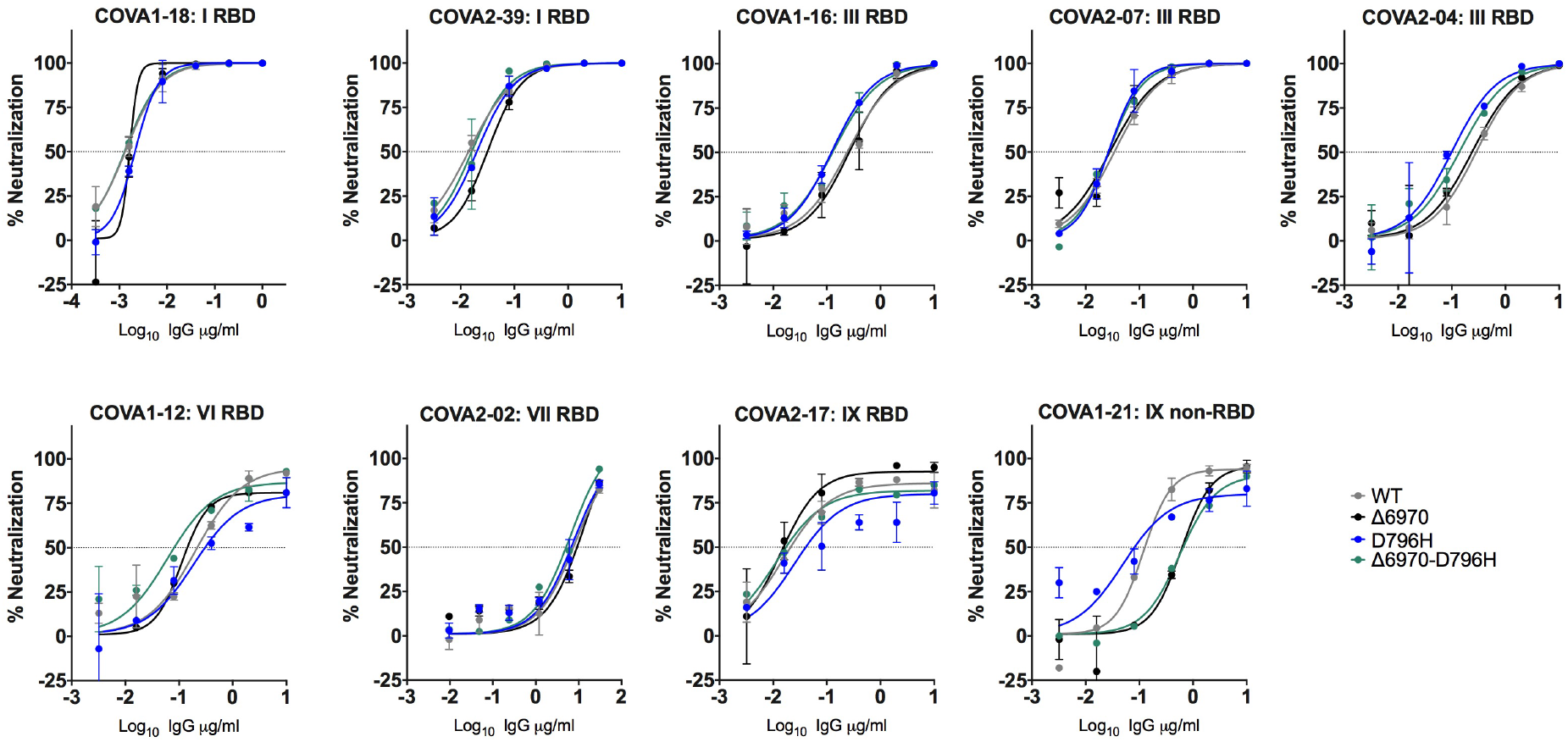
Neutralization potency of a panel of monoclonal antibodies targeting the RBD is not impacted by Spike mutations. Lentivirus pseudotyped with SARS-CoV-2 Spike protein: WT (D614G background), D796H, ΔH69/V70, D796H+ΔH69/V70 were produced in 293T cells and used to infect target Hela cells stably expressing ACE2 in the presence of serial dilutions of indicated monoclonal antibodies. Data are representative of at least two independent experiments. RBD: receptor binding domain. Classes of RBD binding antibodies are indicated based Bouwer et al.

In order to understand the mechanisms that might confer resistance to antibodies we examined a published Spike structure and annotated it with our residues of interest (Figure 8). This analysis showed that **Δ**H69/**Δ**V70 is in a disordered, glycosylated loop at the very tip of the NTD, and therefore could alter binding of antibodies. ΔH69/V70 is close to the binding site of the polyclonal antibodies derived from COV57 plasma^16,17^. D796H is in an exposed loop in S2 (Figure 8) and appears to be in a region frequently targeted by antibodies^18^, despite mutations at position 796 being rare (Supplementary Table 7).

**Figure 8.**
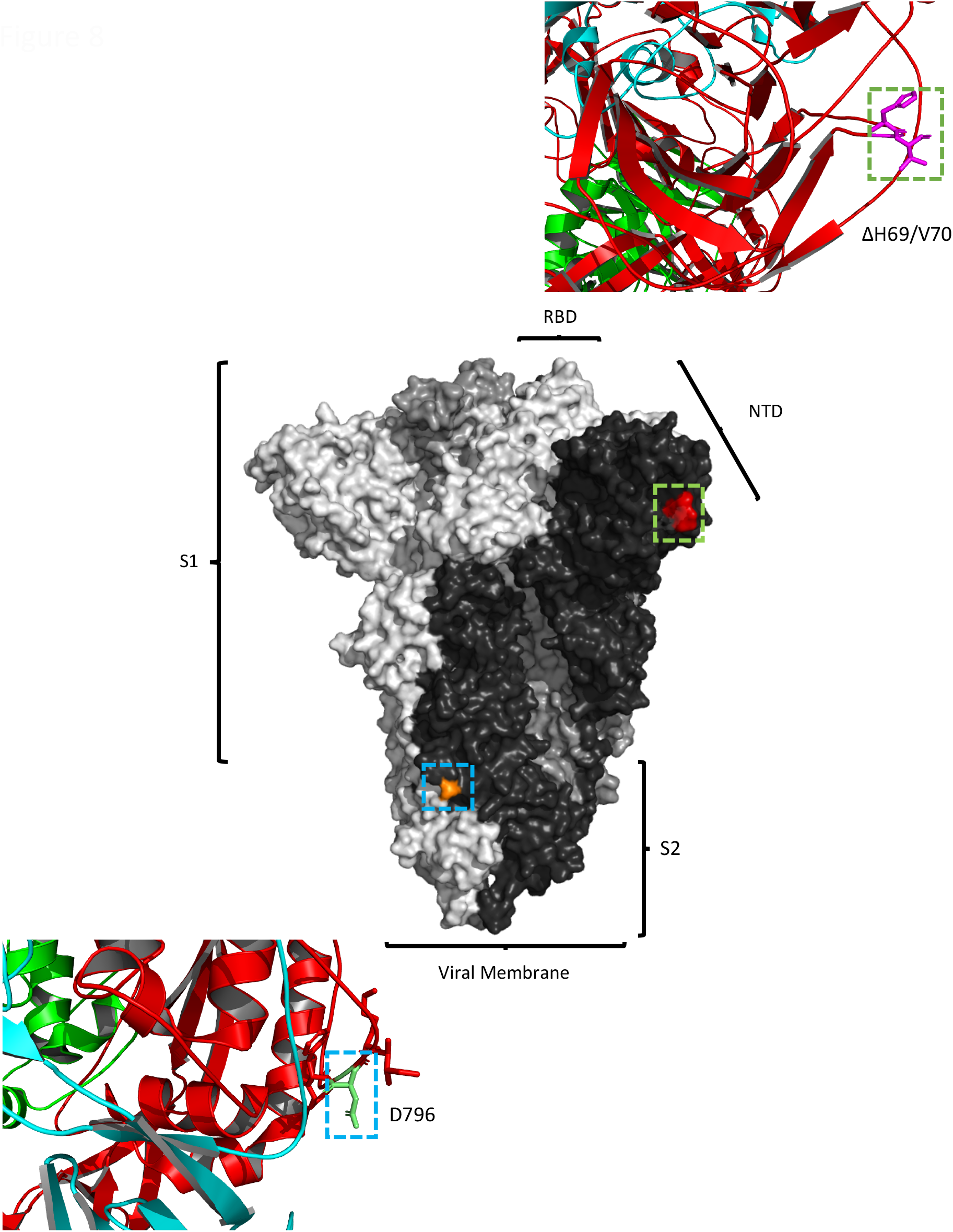
Location of Spike mutations ΔH69/Y70 in S1 and D796H in S2. Amino acid residues H69 and Y70 deleted in the N-terminal domain (red) and D796H in subunit 2 (orange) are highlighted on a SARS-CoV-2 spike trimer (PDB: 6ZGE Wrobek et al., 2020). Each of the three protomers making up the Spike homotrimer are coloured separately in shades of grey (centre). Close-ups of ΔH69/Y70 (above) and D796H (below) are shown in cartoon, stick representation. Both mutations are in exposed loops.

## Discussion

Here we have documented a repeated evolutionary response by SARS-CoV-2 in the presence of antibody therapy during the course of a persistent infection in an immunocompromised host. The observation of potential selection for specific variants coinciding with the presence of antibodies from convalescent plasma is supported by the experimental finding of two-fold reduced susceptibility of these viruses to plasma. Further, we were able to document real-time emergence of a variant **Δ**H69/**Δ**V70 in the NTD of Spike that has been increasing in frequency in Europe and is present in the new UK variant B1.1.7^19^. In this case the emergence of the variant was not the primary reason for treatment failure. However, given that both vaccines and therapeutics are aimed at Spike, our study raises the possibility of virus evasion, particularly in immune suppressed individuals where prolonged viral replication occurs.

Our observations represent a very rare insight, possible due to poor T cell responses and a lack of antibodies in the individual, and an intensive sampling course. Persistent viral replication and the failure of antiviral therapy allowed us to define the viral response to convalescent plasma. Our findings follow those of Choi et al^9^, who reported persistent infection in an immune suppressed individual; they noted significant virus evolution, including NTD deletions and RBD mutations in the absence of SARS-CoV-2 specific antibody therapy. A second paper reported asymptomatic long term shedding with four sequences over 105 days^11^, demonstrated similarly dramatic shifts in genetic composition of the viral population without phenotypic impact. A common finding with our study was the very low neutralisation activity in serum post transfusion of CP with waning over time as expected. Apart from the difference in the outcome of infection (severe, fatal disease versus asymptomatic disease and clearance), critically important differences in our study include: 1. The intensity of sampling and use of both long and short read sequencing to verify variant calls, thereby providing a unique scientific resource for longitudinal population genetic analysis. 2. The close alignment between the genetic composition of the viral population and CP administration, with an experimentally verified variant with reduced susceptibility emerging, falling to low frequency, and then rising again under CP selection. 3. Real time detection of emergence of a variant, **Δ**H69/**Δ**V70, that is increasing in frequency in Europe, and present in the new UK multiply mutated variant B1.1.7.

An interesting observation is that in the two cases of chronic infection highlighted, the Avanzato case where CP was used for asymptomatic shedding^11^ exhibited lower diversification of virus as compared to the fatal Choi et al case where CP was not used^9^. There are clearly a number of factors that could account for these differences, though this highlights the fact that use of CP does not necessarily lead to rapid adaptation. Intriguingly, deletions spanning amino acids 139-145 emerged in both these cases in contrast to the **Δ**H69/**Δ**V70 observed in the present study. Deletion of amino acid 144 and **Δ**H69/**Δ**V70 is observed in the UK lineage B1.1.7^19^, and is therefore of concern.

We have noted in our analysis the potential influence of compartmentalised viral replication upon the sequences recovered in upper respiratory tract samples. Both population genetic and small animal studies have shown a lack of reassortment between influenza viruses within a single host during an infection, suggesting that acute respiratory viral infection may be characterised by spatially distinct viral populations^20,21^. In the analysis of data, it is important to distinguish genetic changes which occur in the primary viral population from apparent changes that arise from the stochastic observation of spatially distinct subpopulations in the host. While the samples we observe on days 93 and 95 of infection are genetically distinct from the others, the remaining samples are consistent with arising from a consistent viral population. We note that Choi et al reported the detection in post-mortem tissue of viral RNA not only in lung tissue, but also in the spleen, liver, and heart^9^. Mixing of virus from different compartments, for example via blood, or movement of secretions from lower to upper respiratory tract, could lead to fluctuations in viral populations at particular sampling sites. Experiments in animal models with sampling of different replication sites could allow a better understanding of SARS-CoV-2 population genetics and enable prediction of escape variants following antibody based therapies.

This is a single case report and therefore limited conclusions can be drawn about generalisability.

In addition to documenting the emergence of SARS-CoV-2 Spike **Δ**H69/**Δ**V70 in vivo, we show that this mutation increases infectivity of the Spike protein in a pseudotyping assay. The deletion was observed contemporaneously with the rare S2 mutation D796H after two separate courses of CP, with other viral populations emerging. D796H, but not **Δ**H69/**Δ**V70, conferred reduction in susceptibility to polyclonal antibodies in the units of CP administered, though we cannot speculate as to their individual impacts on sera from other individuals. Importantly, neither of these mutations alone or in combination affected susceptibility of virus to a set of RBD-targeting monoclonal antibodies. The reduced sensitivity of **Δ**H69/**Δ**V70 to a non RBD binding antibody may hint at a role in antigen recognition, for example to polyclonal sera from COV-57.

The effects of CP on virus evolution seen here are unlikely to apply in immune competent hosts where viral diversity is likely to be lower due to better immune control. Our data highlight that infection control measures may need to be tailored to the needs of immunocompromised patients and also caution in interpretation of CDC guidelines that recommend 20 days as the upper limit of infection prevention precautions in immune compromised patients who are afebrile^22^. Due to the difficulty with culturing clinical isolates, use of surrogates are warranted^23^. However, where detection of ongoing viral evolution is possible, this serves as a clear proxy for the existence of infectious virus. In our case we detected environmental contamination whilst in a single occupancy room and the patient was moved to a negative-pressure high air-change infectious disease isolation room.

Clinical efficacy of convalescent plasma in severe COVID-19 has not been demonstrated^24^, and its use in different stages of infection and disease remains experimental; as such, we suggest that it should be reserved for use within clinical trials, with rigorous monitoring of clinical and virological parameters. The data from this single case report might warrant caution in use of convalescent plasma in patients with immune suppression of both T cell and B cell arms; in such cases, the antibodies administered have little support from cytotoxic T cells, thereby reducing chances of clearance and theoretically raising the potential for escape mutations. Whilst we await further data, where clinical trial enrolment is not possible, convalescent plasma administered for clinical need in immune suppression should ideally only be considered as part of observational studies, undertaken preferably in single occupancy rooms with enhanced infection control precautions, including SARS-CoV-2 environmental sampling and real-time sequencing. Understanding of viral dynamics and characterisation of viral evolution in response to different selection pressures in the immunocompromised host is necessary not only for improved patient management but also for public health benefit.

## Ethics

The study was approved by the East of England – Cambridge Central Research Ethics Committee (17/EE/0025). Written informed consent was obtained from both the patient and family. Additional controls with COVID-19 were enrolled to the NIHR BioResource Centre Cambridge under ethics review board (17/EE/0025).

## Acknowledgements

We are immensely grateful to the patient and his family. We would also like to thank the staff at CUH and the NIHR Cambridge Clinical Research Facility. We would like to thank Dr Ruthiran Kugathasan and Professor Wendy Barclay for helpful discussions and Dr Martin Curran, Dr William Hamilton, and Dr. Dominic Sparkes. We would like to thank Prof Andres Floto and Prof Ferdia Gallagher. We thank Dr James Voss for the kind gift of HeLa cells stably expressing ACE2. COG-UK is supported by funding from the Medical Research Council (MRC) part of UK Research & Innovation (UKRI), the National Institute of Health Research (NIHR) and Genome Research Limited, operating as the Wellcome Sanger Institute. RKG is supported by a Wellcome Trust Senior Fellowship in Clinical Science (WT108082AIA). LEM is supported by a Medical Research Council Career Development Award (MR/R008698/1). SAK is supported by the Bill and Melinda Gates Foundation via PANGEA grant: OPP1175094. DAC is supported by a Wellcome Trust Clinical PhD Research Fellowship. CJRI acknowledges MRC funding (ref: MC_UU_00002/11). This research was supported by the National Institute for Health Research (NIHR) Cambridge Biomedical Research Centre, the Cambridge Clinical Trials Unit (CCTU) and by the UCL Coronavirus Response Fund and made possible through generous donations from UCL’s supporters, alumni, and friends (LEM). JAGB is supported by the Medical Research Council (MC_UP_1201/16). IG is a Wellcome Senior Fellow and supported by the Wellcome Trust (207498/Z/17/Z). DDP is supported by NIH GM083127.

## Author contributions

Conceived study: RKG, SAK, DAC, AS, TG, EGK

Designed experiments: RKG, SAK, DAC, LEM, JAGB, EGK, AC, NT, AC, CS, RD, RG, DDP

Performed experiments: SAK, DAC, LEM, RD, CRS, AJ, IATMF, KS, TG, CJRI, BB, JS, MJvG, LGC, GBM

Interpreted data: RKG, SAK, DAC, PM, LEM, JAGB, PM, SG, KS, TG, JB, KGCS, IG, CJRI, JAGB, IUL, DR, JS, BB, RAG. DDP, RD, LCG, GBM

## Methods

### Clinical Sample Collection and Next generation sequencing

Serial samples were collected from the patient periodically from the lower respiratory tract (sputum or endotracheal aspirate), upper respiratory tract (throat and nasal swab), and from stool. Nucleic acid extraction was done from 500µl of sample with a dilution of MS2 bacteriophage to act as an internal control, using the easyMAG platform (Biomerieux, Marcy-l’Étoile) according to the manufacturers’ instructions. All samples were tested for presence of SARS-CoV-2 with a validated one-step RT q-PCR assay developed in conjunction with the Public Health England Clinical Microbiology ^25^. Amplification reaction were all performed on a Rotorgene™ PCR instrument. Samples which generated a CT of ≤36 were considered to be positive.

Sera from recovered patients in the COVIDx study^26^ were used for testing of neutralisation activity by SARS-CoV-2 mutants.

#### SARS-CoV-2 serology by multiplex particle-based flow cytometry (Luminex)

Recombinant SARS-CoV-2 N, S and RBD were covalently coupled to distinct carboxylated bead sets (Luminex; Netherlands) to form a 3-plex and analyzed as previously described (Xiong et al. 2020). Specific binding was reported as mean fluorescence intensities (MFI).

#### Whole blood T cell and innate stimulation assay

Whole blood was diluted 1:5 in RPMI into 96-well F plates (Corning) and activated by single stimulation with phytohemagglutinin (PHA; 10 μg/ml; Sigma-Aldrich), or LPS (1 μg/ml, List Biochemicals) or by co-stimulating with anti-CD3 (MEM57, Abcam, 200 ng/ml,) and IL-2 (Immunotools, 1430U/ml,). Supernatants were taken after 24 hours. Levels (pg/ml) are shown for IFNg, IL17, IL2, TNFa, IL6, IL1b and IL10. Cytokines were measured by multiplexed particle based Flow cytometry on a Luminex analyzer (Bio-Plex, Bio-Rad, UK) using an R&D Systems custom kit (R&D Systems, UK).

For viral genomic sequencing, total RNA was extracted from samples as described. Samples were sequenced using MinION flow cells version 9.4.1 (Oxford Nanopore Technologies) following the ARTICnetwork V3 protocol (https://dx.doi.org/10.17504/protocols.io.bbmuik6w) and BAM files assembled using the ARTICnetwork assembly pipeline (https://artic.network/ncov-2019/ncov2019-bioinformatics-sop.html). A representative set of 10 sequences were selected and also sequenced using the Illumina MiSeq platform. Amplicons were diluted to 2 ng/µl and 25 µl (50 ng) were used as input for each library preparation reaction. The library preparation used KAPA Hyper Prep kit (Roche) according to manufacturer’s instructions. Briefly, amplicons were end-repaired and had A-overhang added; these were then ligated with 15mM of NEXTflex DNA Barcodes (Bio Scientific, Texas, USA). Post-ligation products were cleaned using AMPure beads and eluted in 25 µl. Then, 20 µl were used for library amplification by 5 cycles of PCR. For the negative controls, 1ng was used for ligation-based library preparation. All libraries were assayed using TapeStation (Agilent Technologies, California, USA) to assess fragment size and quantified by QPCR. All libraries were then pooled in equimolar accordingly. Libraries were loaded at 15nM and spiked in 5% PhiX (Illumina, California, USA) and sequenced on one MiSeq 500 cycle using a Miseq Nano v2 with 2x 250 paired-end sequencing. A minimum of ten reads were required for a variant call.

### Bioinformatics Processes

For long-read sequencing, genomes were assembled with reference-based assembly and a curated bioinformatics pipeline with 20x minimum coverage across the whole-genome ^27^. For short-read sequencing, FASTQs were downloaded, poor-quality reads were identified and removed, and both Illumina and PHiX adapters were removed using TrimGalore v0.6.6 ^28^. Trimmed paired-end reads were mapped to the National Center for Biotechnology Information SARS-CoV-2 reference sequence MN908947.3 using MiniMap2-2.17 with arguments -ax and sr ^29^. BAM files were then sorted and indexed with samtools v1.11 and PCR optical duplicates removed using Picard (http://broadinstitute.github.io/picard). A consensus sequences of nucleic acids with a minimum whole-genome coverage of at least 20× were generated with BCFtools using a 0% majority threshold.

### Single Genome Amplification and sequencing

Viral RNA extracts were reverse transcribed from each sample to sufficiently capture the diversity of the viral population without introducing resampling bias. SuperScript IV (Thermofisher Scientific) and the gene specific primers were used for reverse transcription. Template RNA was degraded with RNAse H (Thermofisher Scientific). All primers used were ‘in-house’ primers designed using the multiple sequence alignment of the patient’s consensus NGS sequences. Partial Spike (amino acids 21-800) was amplified as 1 continuous length of DNA (Spike ∼ 1.8 kb) by nested PCR. Terminally diluted cDNA was PCR-amplified using Platinum^®^ Taq DNA Polymerase High Fidelity (Invitrogen, Carlsbad, CA) so that 30% of reactions were positive^30^. By Poisson statistics, sequences were deemed ≥80% likely to be derived from HIV-1 single genomes. We obtained between 20–60 single genomes at each sample time point to achieve 90% confidence of detecting variants present at ≥8% of the viral population in vivo^31,32^. Partial spike amplicons obtained from terminal dilution PCR amplification were Sanger sequenced to form a contiguous sequence using another set of 8 in-house primers. Sanger sequencing was provided by Genewiz UK and manual sequence editing was performed using DNA Dynamo software (Blue Tractor Software Ltd, UK).

### Phylogenetic Analysis

All available full-genome SARS-CoV-2 sequences were downloaded from the GISAID database (http://gisaid.org/) ^33^ on 16^th^ December. Duplicate and low-quality sequences (>5% N regions) were removed, leaving a dataset of 212,297 sequences with a length of >29,000bp. All sequences were sorted by name and only sequences sequenced with United Kingdom / England identifiers were retained. From this dataset, sequecnes were de-duplicated and where background sequences were required in figures, randomly subsampled using seqtk (https://github.com/lh3/seqtk). All sequences were aligned to the SARS-CoV-2 reference strain MN908947.3, using MAFFT v7.475 with automatic flavour selection^34^. Major SARS-CoV-2 clade memberships were assigned to all sequences using both the Nextclade server v0.9 (https://clades.nextstrain.org/) and Phylogenetic Assignment Of Named Global Outbreak Lineages (pangolin)^35^.

Maximum likelihood phylogenetic trees were produced using the above curated dataset using IQ-TREE v2.1.2^36^. Evolutionary model selection for trees were inferred using ModelFinder^37^ and trees were estimated using the GTR+F+I model with 1000 ultrafast bootstrap replicates^38^. All trees were visualised with Figtree v.1.4.4 (http://tree.bio.ed.ac.uk/software/figtree/), rooted on the SARS-CoV-2 reference sequence and nodes arranged in descending order. Nodes with bootstraps values of <50 were collapsed using an in-house script.

### In-depth allele frequency variant calling

The SAMFIRE package^39^ was used to call allele frequency trajectories from BAM file data. Reads were included in this analysis if they had a median PHRED score of at least 30, trimming the ends of reads to achieve this if necessary. Nucleotides were then filtered to have a PHRED score of at least 30; reads with fewer than 30 such reads were discarded. Distances between sequences, accounting for low-frequency variant information, was also conducted using SAMFIRE. The sequence distance metric, described in an earlier paper ^40^, combines allele frequencies across the whole genome. Where L is the length of the genome, we define q(t) as a 4 x L element vector describing the frequencies of each of the nucleotides A, C, G, and T at each locus in the viral genome sampled at time t. For any given locus i in the genome we calculate the change in allele frequencies between the times t1 and t2 via a generalisation of the Hamming distance

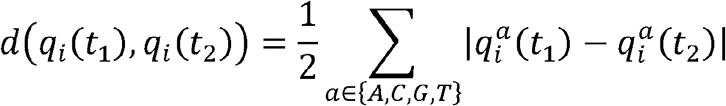

where the vertical lines indicate the absolute value of the difference. These statistics were then combined across the genome to generate the pairwise sequence distance metric

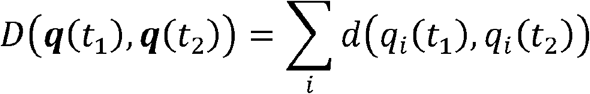

The Mathematica software package was to conduct a regression analysis of pairwise sequence distances against time, leading to an estimate of a mean rate of within-host sequence evolution. In contrast to the phylogenetic analysis, this approach assumed the samples collected on days 93 and 95 to arise via stochastic emission from a spatially separated subpopulation within the host, leading to a lower inferred rate of viral evolution for the bulk of the viral population.

### Western blot analysis

Forty-eight hours after transfection of cells with plasmid preparations, the culture supernatant was harvested and passed through a 0.45-μm-pore-size filter to remove cellular debris. The filtrate was centrifuged at 15,000 rpm for 120 min to pellet virions. The pelleted virions were lysed in Laemmli reducing buffer (1 M Tris-HCl [pH 6.8], SDS, 100% glycerol, β-mercaptoethanol, and bromophenol blue). Pelleted virions were subjected to electrophoresis on SDS–4 to 12% bis-Tris protein gels (Thermo Fisher Scientific) under reducing conditions. This was followed by electroblotting onto polyvinylidene difluoride (PVDF) membranes. The SARSCOV-2 Spike proteins were visualized by a ChemiDoc^®^ MP imaging system (Biorad) using anti-Spike S2 (Invitrogen) and anti-p24 Gag antibodies.

### Recombination Detection

All sequences were tested for potential recombination, as this would impact on evolutionary estimates. Potential recombination events were explored with nine algorithms (RDP, MaxChi, SisScan, GeneConv, Bootscan, PhylPro, Chimera, LARD and 3SEQ), implemented in RDP5 with default settings ^41^. To corroborate any findings, ClonalFrameML v1.12 ^42^ was also used to infer recombination breakpoints. Neither programs indicated evidence of recombination in our data.

### Structural Viewing

The Pymol Molecular Graphics System v2.4.0 (https://github.com/schrodinger/pymol-open-source/releases) was used to map the location of the four spike mutations of interested onto a SARS-CoV-2 spike structure visualised by Wrobel et al (PDB: 6ZGE)^43^.

### Testing of convalescent plasma for antibody titres

The Anti-SARS-CoV-2 ELISA (IgG) assay used to test CP for antibody titres was Euroimmun Medizinische Labordiagnostika AG. This indirect ELISA based assay uses a recombinant structural spike 1 (S1) protein of SARS-CoV-2 expressed in the human cell line HEK 29 for the detection of SARS-CoV2 IgG.

### Generation of Spike mutants

Amino acid substitutions were introduced into the D614G pCDNA_SARS-CoV-2_Spike plasmid as previously described^44^ using the QuikChange Lightening Site-Directed Mutagenesis kit, following the manufacturer’s instructions (Agilent Technologies, Inc., Santa Clara, CA).

### Pseudotype virus preparation

Viral vectors were prepared by transfection of 293T cells by using Fugene HD transfection reagent (Promega). 293T cells were transfected with a mixture of 11ul of Fugene HD, 1µg of pCDNAΔ19Spike-HA, 1ug of p8.91 HIV-1 gag-pol expression vector^45,46^, and 1.5µg of pCSFLW (expressing the firefly luciferase reporter gene with the HIV-1 packaging signal). Viral supernatant was collected at 48 and 72h after transfection, filtered through 0.45um filter and stored at -80°C. The 50% tissue culture infectious dose (TCID50) of SARS-CoV-2 pseudovirus was determined using Steady-Glo Luciferase assay system (Promega).

### Standardisation of virus input by SYBR Green-based product-enhanced PCR assay (SG-PERT)

The reverse transcriptase activity of virus preparations was determined by qPCR using a SYBR Green-based product-enhanced PCR assay (SG-PERT) as previously described^47^.

Briefly, 10-fold dilutions of virus supernatant were lysed in a 1:1 ratio in a 2x lysis solution (made up of 40% glycerol v/v 0.25% Trition X-100 v/v 100mM KCl, RNase inhibitor 0.8 U/ml, TrisHCL 100mM, buffered to pH7.4) for 10 minutes at room temperature.

12µl of each sample lysate was added to thirteen 13µl of a SYBR Green master mix (containing 0.5µM of MS2-RNA Fwd and Rev primers, 3.5pmol/ml of MS2-RNA, and 0.125U/µl of Ribolock RNAse inhibitor and cycled in a QuantStudio. Relative amounts of reverse transcriptase activity were determined as the rate of transcription of bacteriophage MS2 RNA, with absolute RT activity calculated by comparing the relative amounts of RT to an RT standard of known activity.

### Serum/plasma pseudotype neutralization assay

Spike pseudotype assays have been shown to have similar characteristics as neutralisation testing using fully infectious wild type SARS-CoV-2^14^.Virus neutralisation assays were performed on 293T cell transiently transfected with ACE2 and TMPRSS2 using SARS-CoV-2 Spike pseudotyped virus expressing luciferase^48^. Pseudotyped virus was incubated with serial dilution of heat inactivated human serum samples or convalescent plasma in duplicate for 1h at 37°C. Virus and cell only controls were also included. Then, freshly trypsinized 293T ACE2/TMPRSS2 expressing cells were added to each well. Following 48h incubation in a 5% CO2 environment at 37°C, the luminescence was measured using Steady-Glo Luciferase assay system (Promega).

### mAb pseudotype neutralisation assay

Virus neutralisation assays were performed on HeLa cells stably expressing ACE2 and using SARS-CoV-2 Spike pseudotyped virus expressing luciferase as previously described^49^. Pseudotyped virus was incubated with serial dilution of purified mAbs^15^ in duplicate for 1h at 37°C. Then, freshly trypsinized HeLa ACE2-expressing cells were added to each well. Following 48h incubation in a 5% CO_2_ environment at 37°C, the luminescence was measured using Bright-Glo Luciferase assay system (Promega) and neutralization calculated relative to virus only controls. IC50 values were calculated in GraphPad Prism.

## Data Availability

Long-read sequencing data that support the findings of this study have been deposited in the NCBI SRA database with the accession codes SAMN16976824 - SAMN16976846 under BioProject PRJNA682013 (https://www.ncbi.nlm.nih.gov/bioproject/PRJNA682013). Short reads and data used to construct figures were deposited at https://github.com/Steven-Kemp/sequence_files.

---

**Supplemental Table 1:**
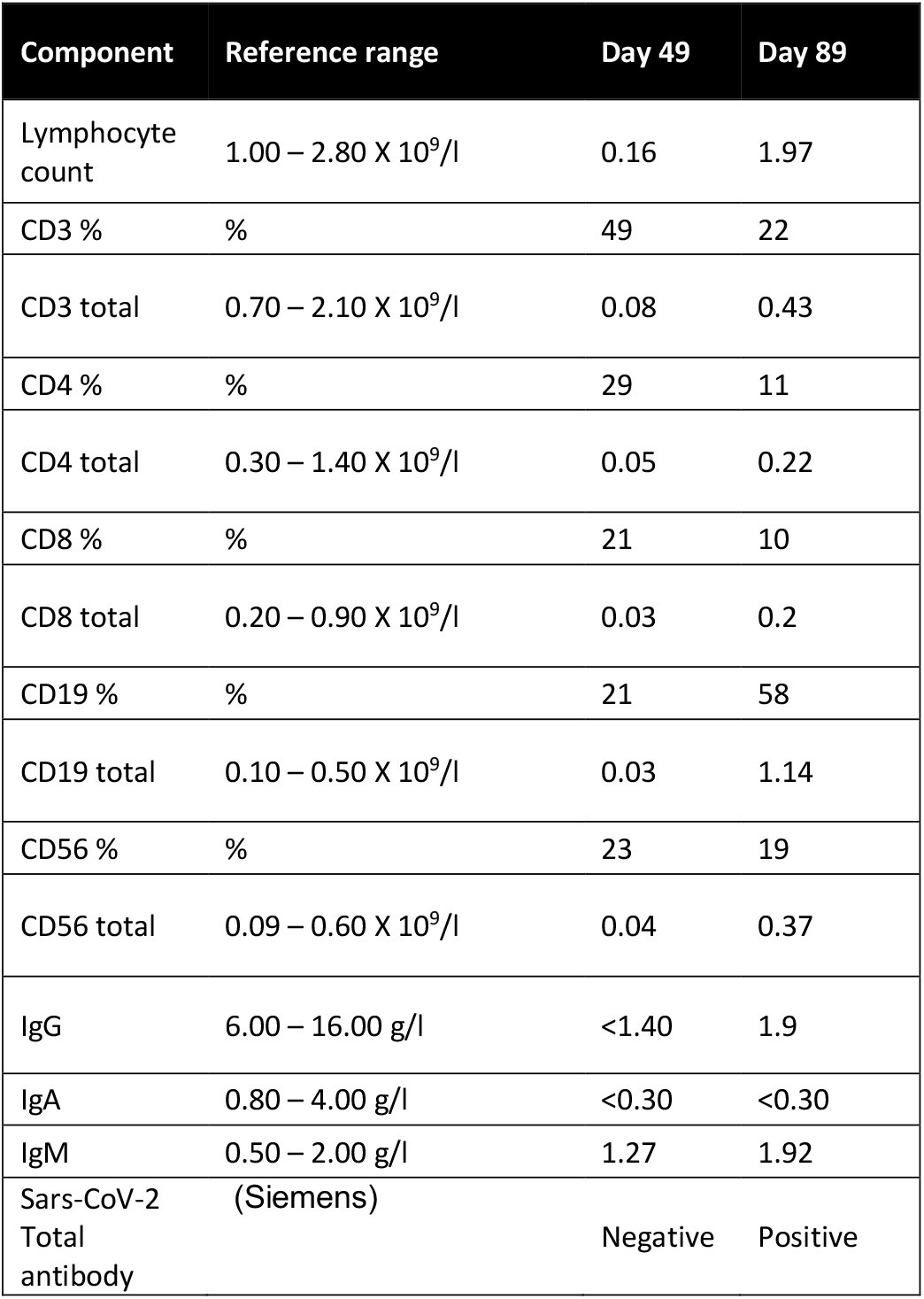
Immunological parameters of patient pre- and post-receipt of convalescent plasma (CP) demonstrating profound lymphopenia with reduction in total serum IgG and IgA. The patient continued to have normal serum IgM which either reflects his underlying lymphoproliferative state or ongoing infection. Following receipt of CP the presence of SARS-CoV-2 antibodies is evident.

**Supplementary Table 2.**
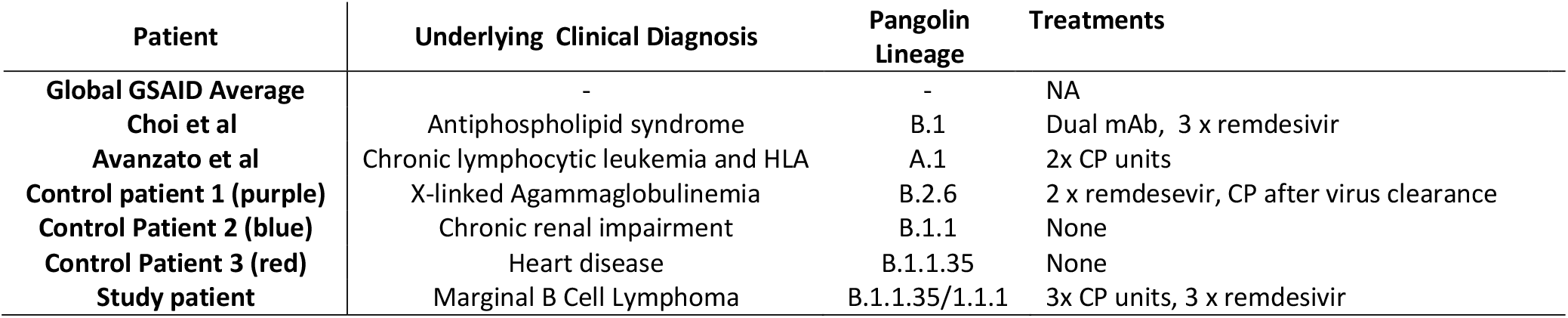
Characteristics of genomes used in the phylogenetic analysis in Figure 2B. Pangolin lineages were identified using Pangolin COVID-19 Lineage Assigner v2.0.8 (https://pangolin.cog-uk.io/).. CP-convalescent plasma; mAb – monoclonal antibody.

**Supplementary Table 3:**
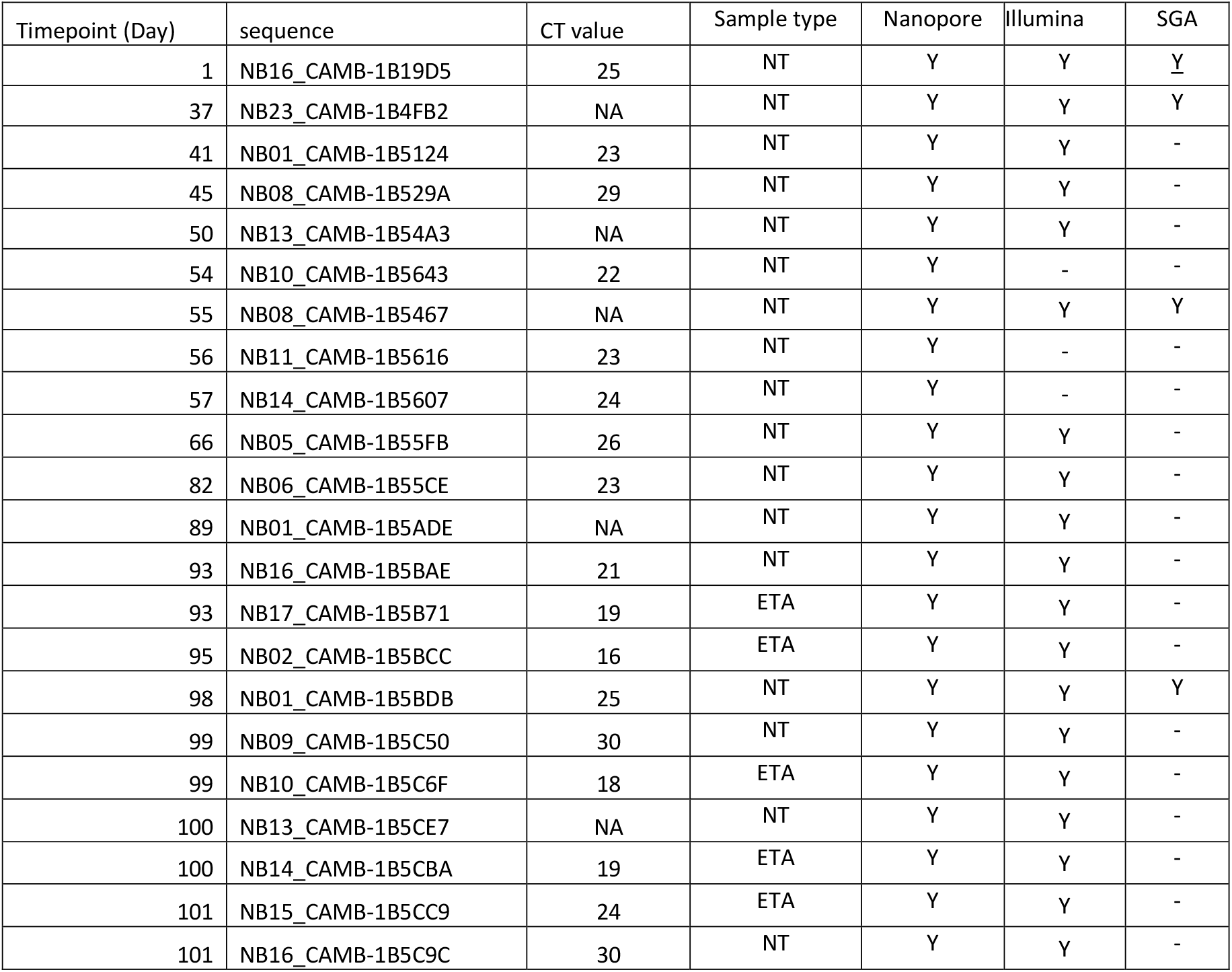
Samples and sequencing methods. Timepoint indicates the day since 1^st^ positive qPCR for SARS-COV-2 that the sample was taken. CT values are reported where available. Y-Yes, - Not done, NA not available

**Supplementary Table 4.**
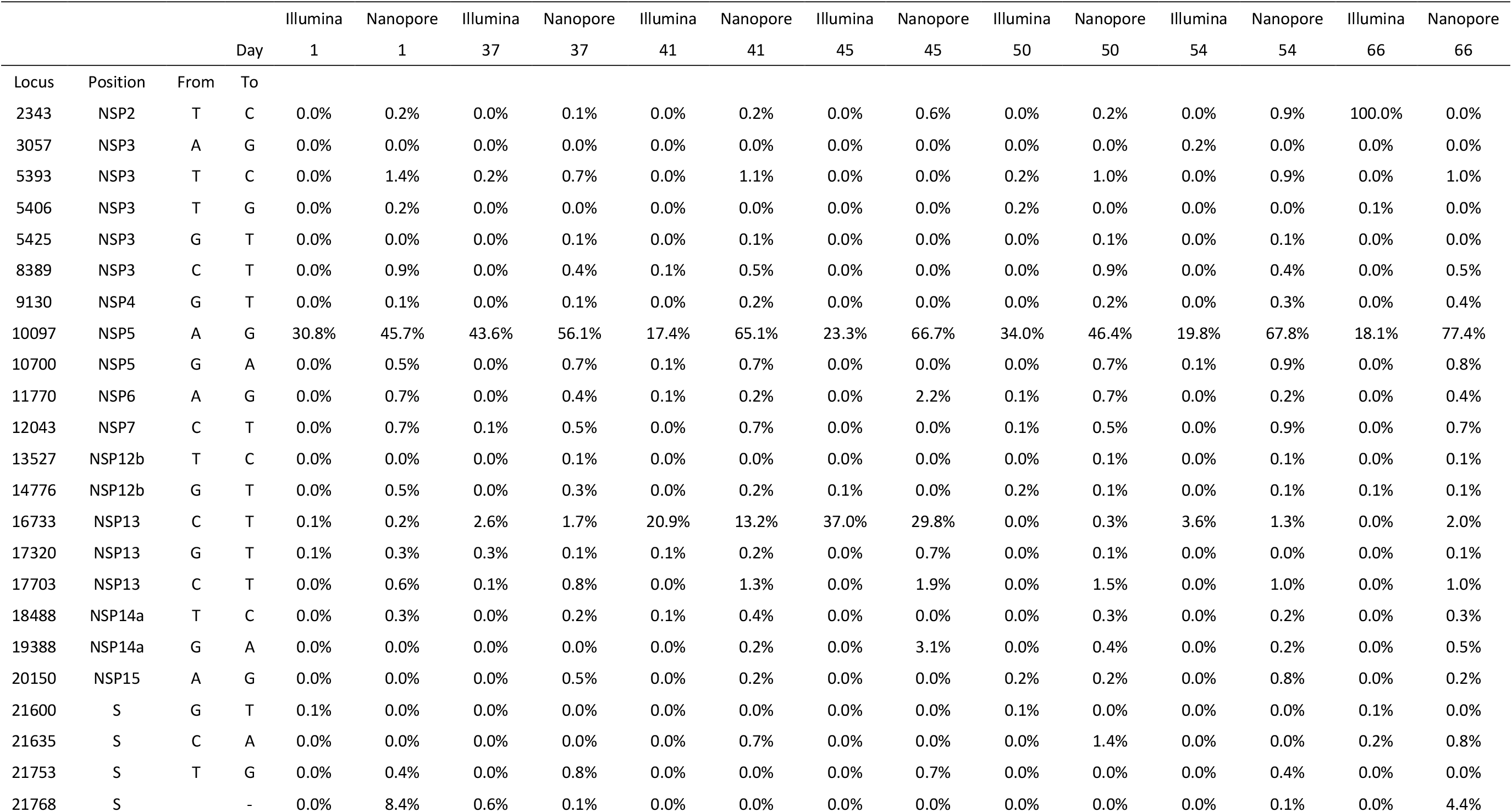
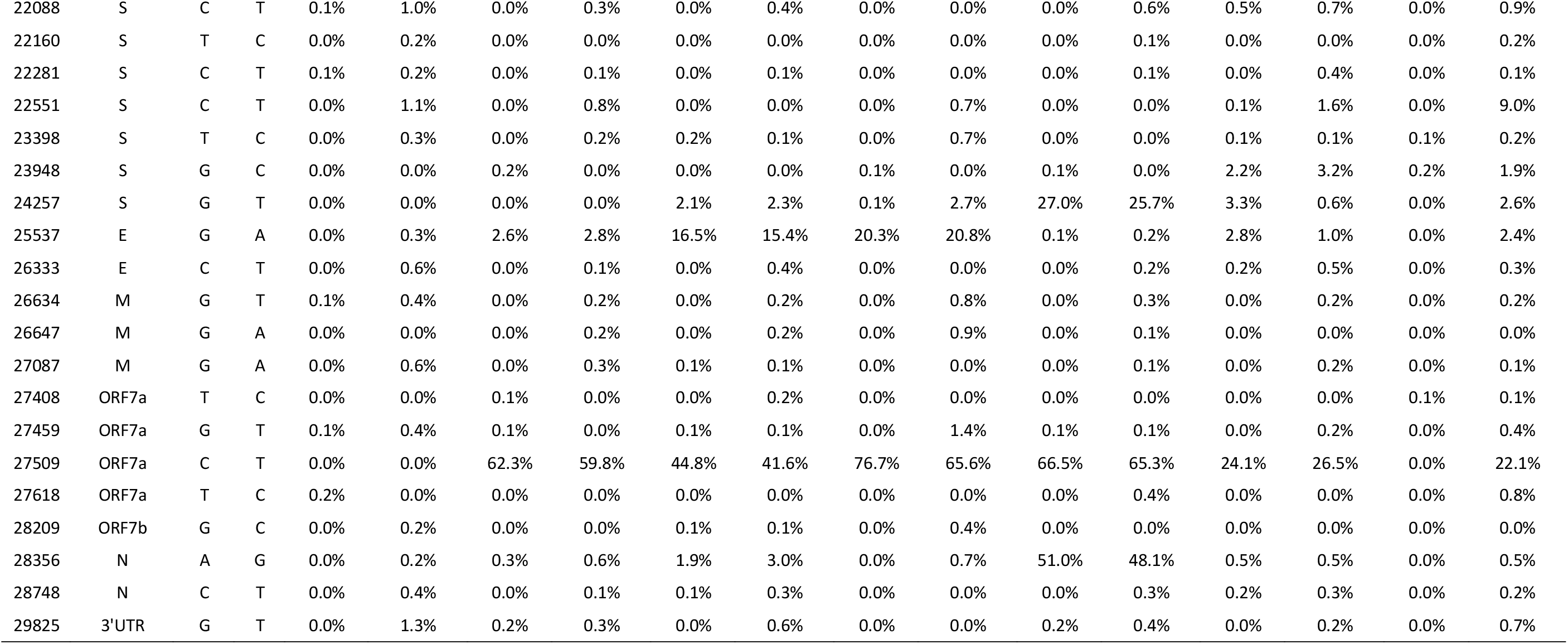
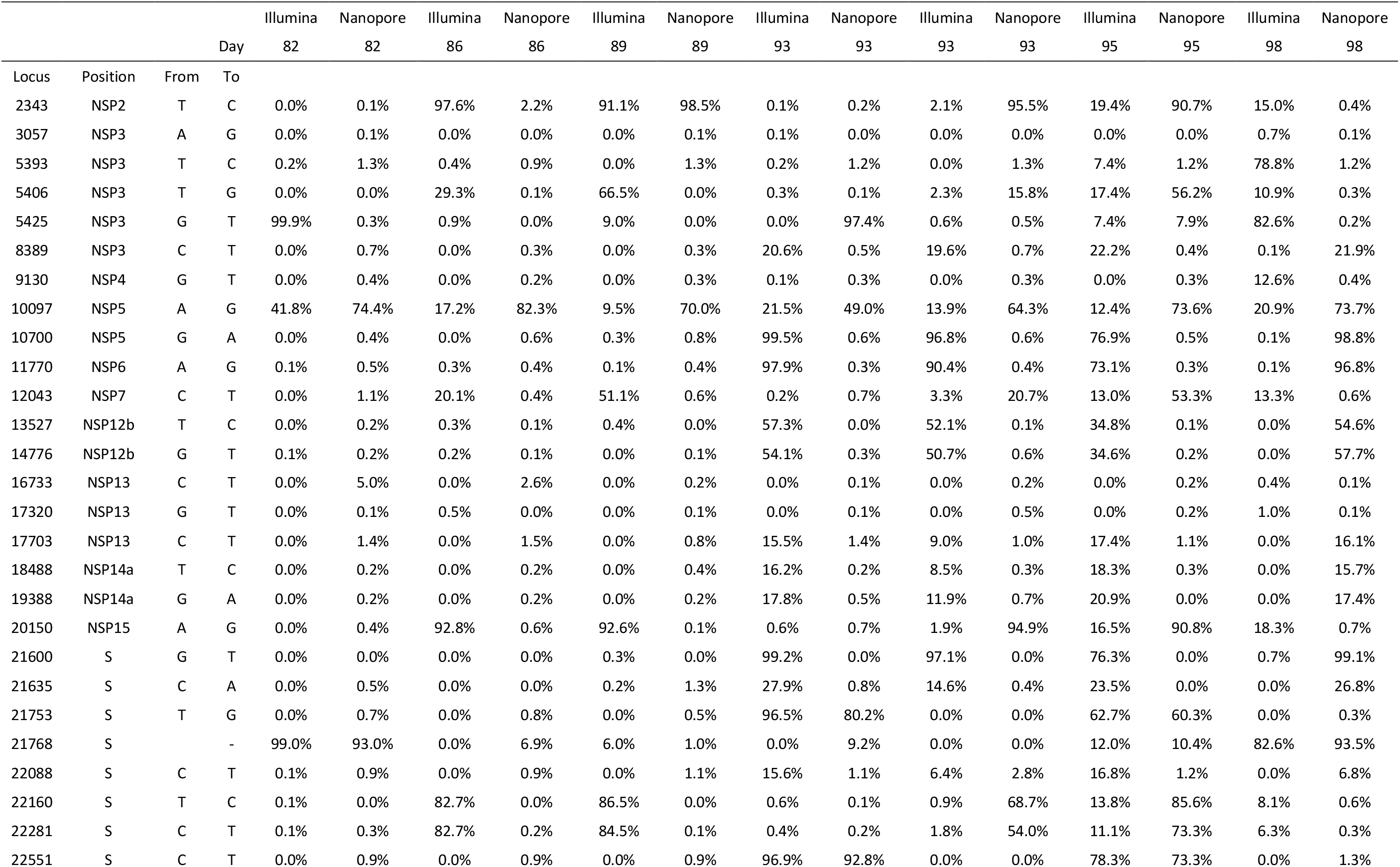
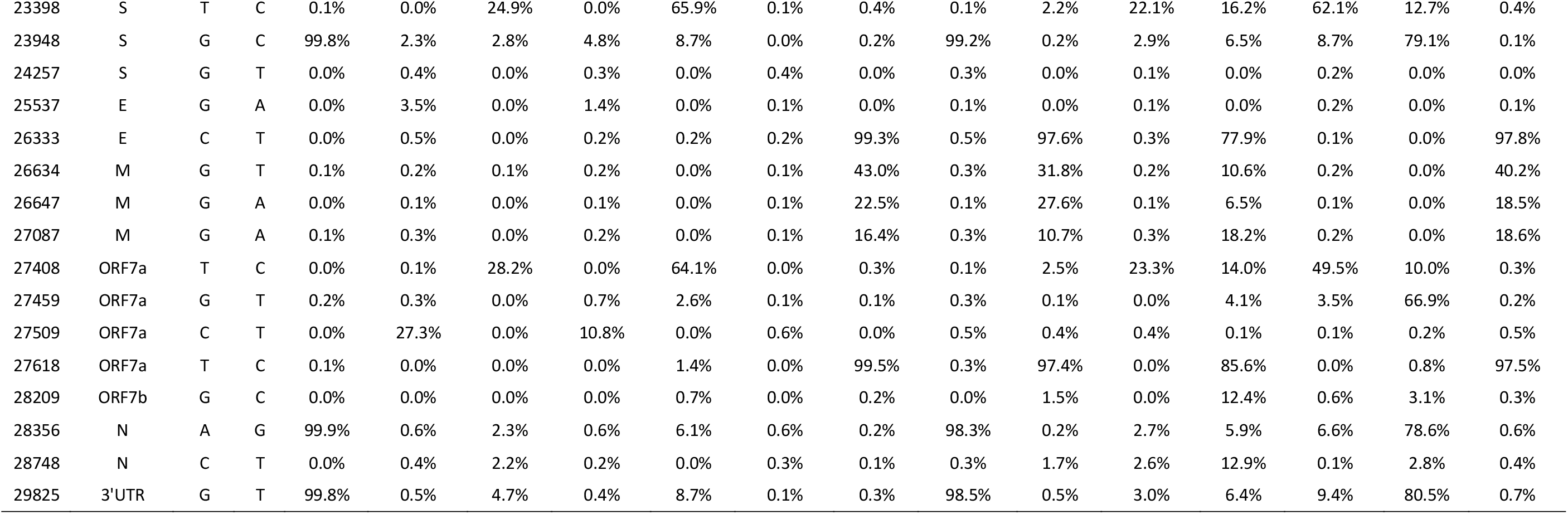
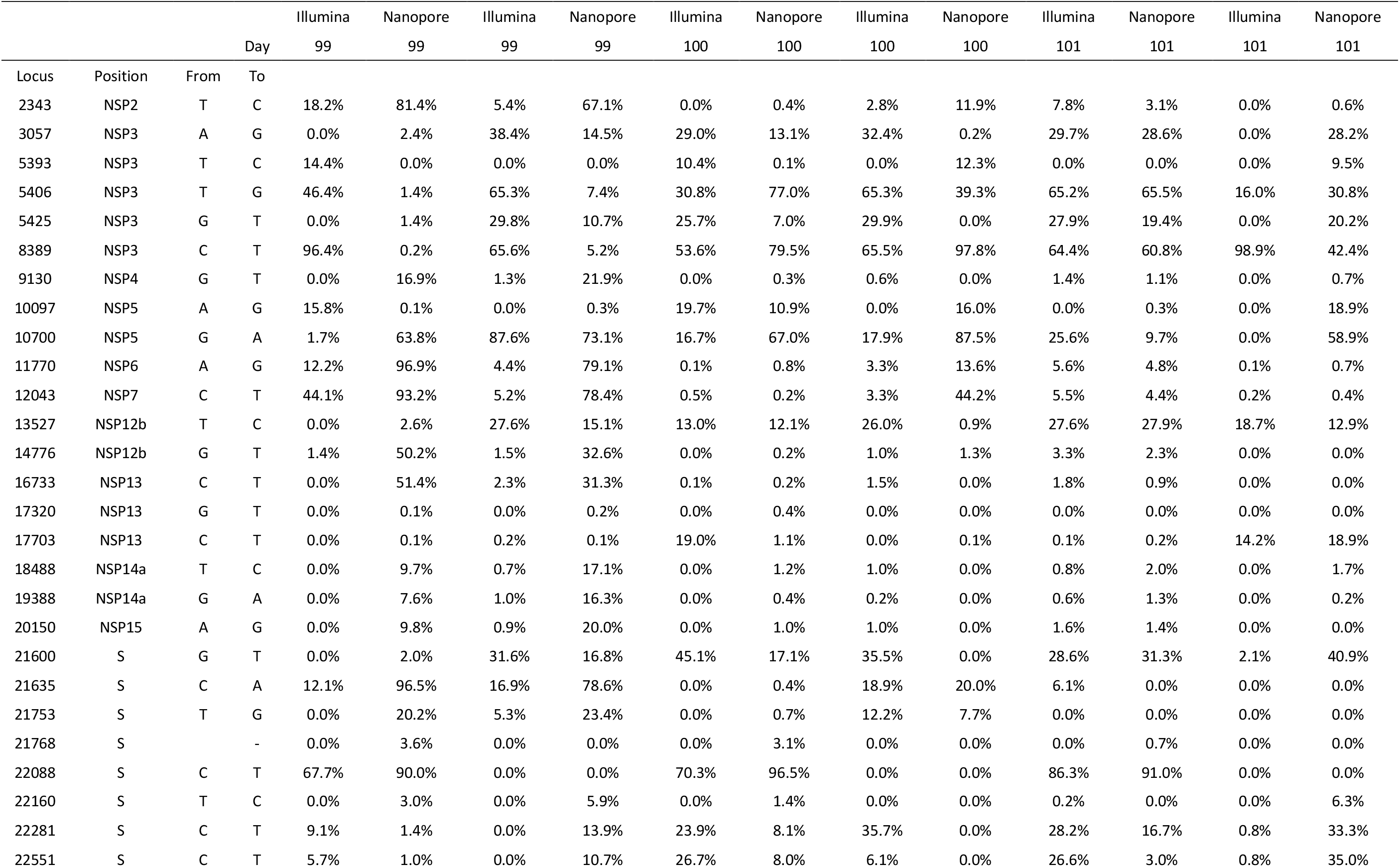
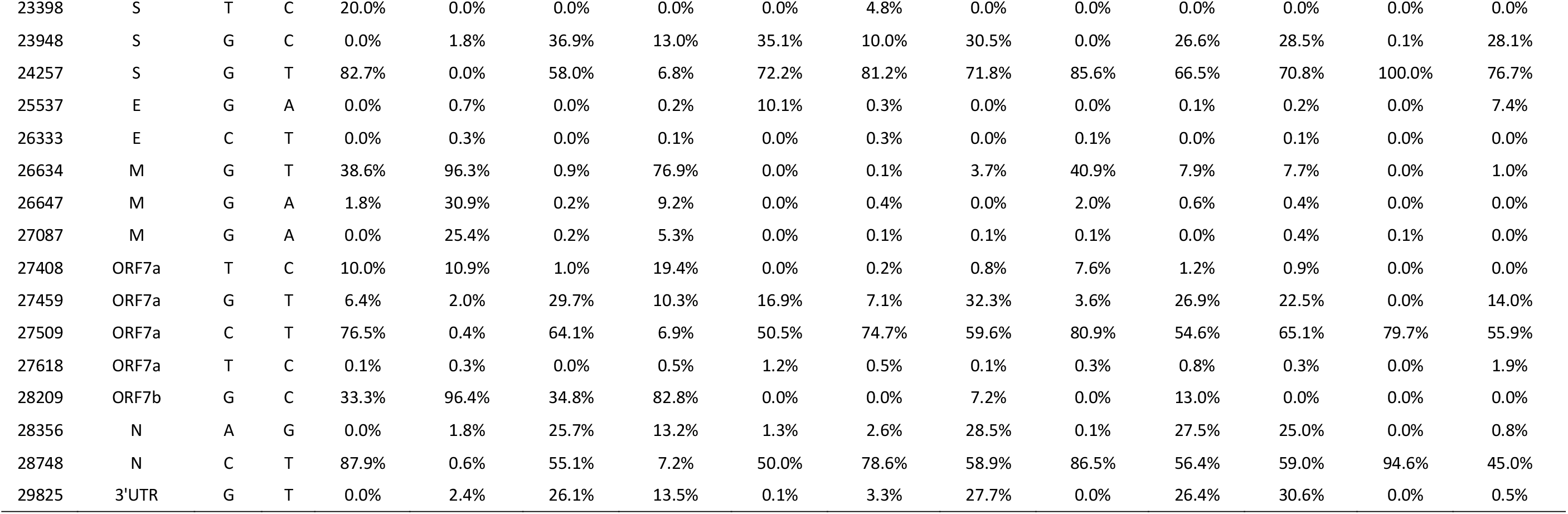
Prevalence of selected Spike glycoprotein mutations at sequential time points and sequencing depth (number of reads covering the amino acid position) measured by both short-read (Illumina) and long-read (Oxford Nanopore) methods. There was low coverage of S:P330S and S:D796H in the final three timepoints, measured by both short- and long-read methods.

**Supplementary Table 5:**
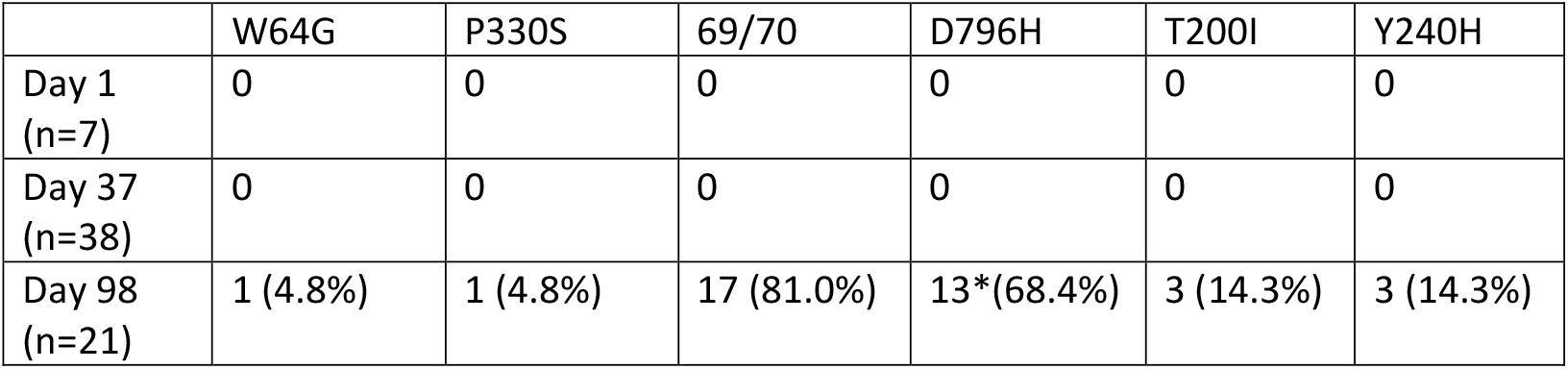
Single genome sequencing (SGS) data from respiratory samples at indicated days. Indicated are the number of single genomes obtained at each time point with the mutations of interest (identified by deep sequencing). *denominator is 19 as for 2 samples the primer reads were poor quality at amino acid 796 at day 98.

**Supplementary Table 6:**
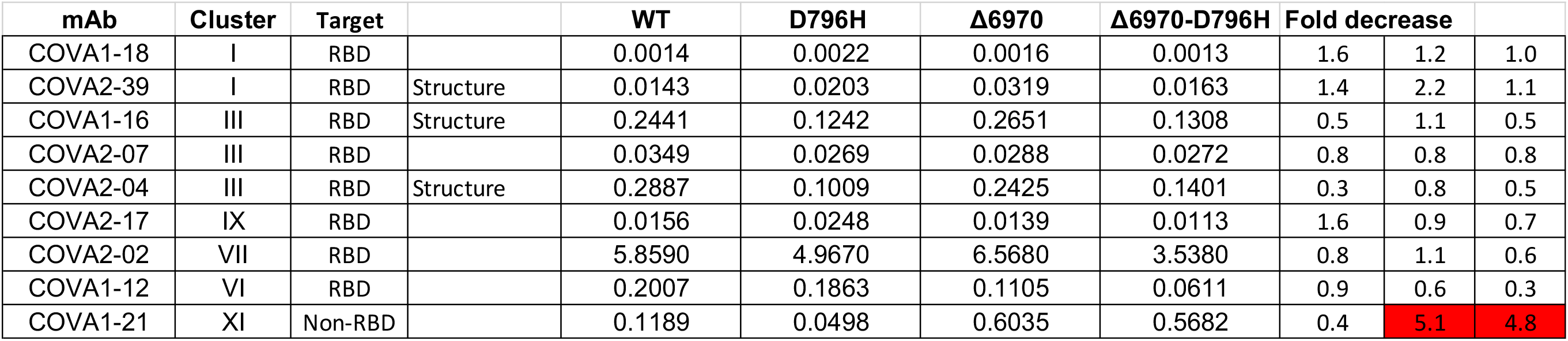
Neutralisation of mutants by *Seven RBD-specific mAbs* (*from Bauwer et al. in Figure 7*). Clusters II, V contain only non-neutralising mAbs, smaller neutralising mAb clusters IV (n=2) and X (n=1) were not tested. Red indicates significant fold changes.

**Supplementary Table 7.**
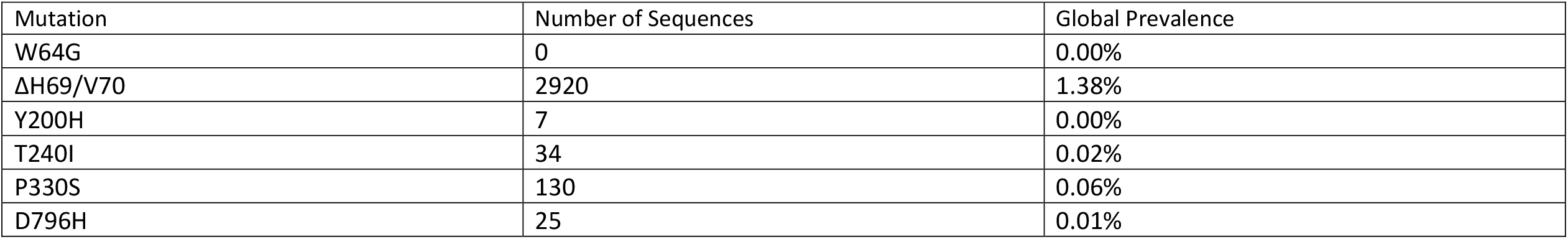
Global prevalence of selected spike mutations detailed in this paper. All high coverage sequences were downloaded from the GISAID database on 11^th^ November and aligned using MAFFT. The global prevalence of each of the six spike mutations W64G, ΔH69/V70, Y200H, T240I, P330S and D796H were assessed by viewing the multiple sequence alignment in AliView, sorting by the column of interest, and counting the number of mutations.

**Supplementary Figure 1:**
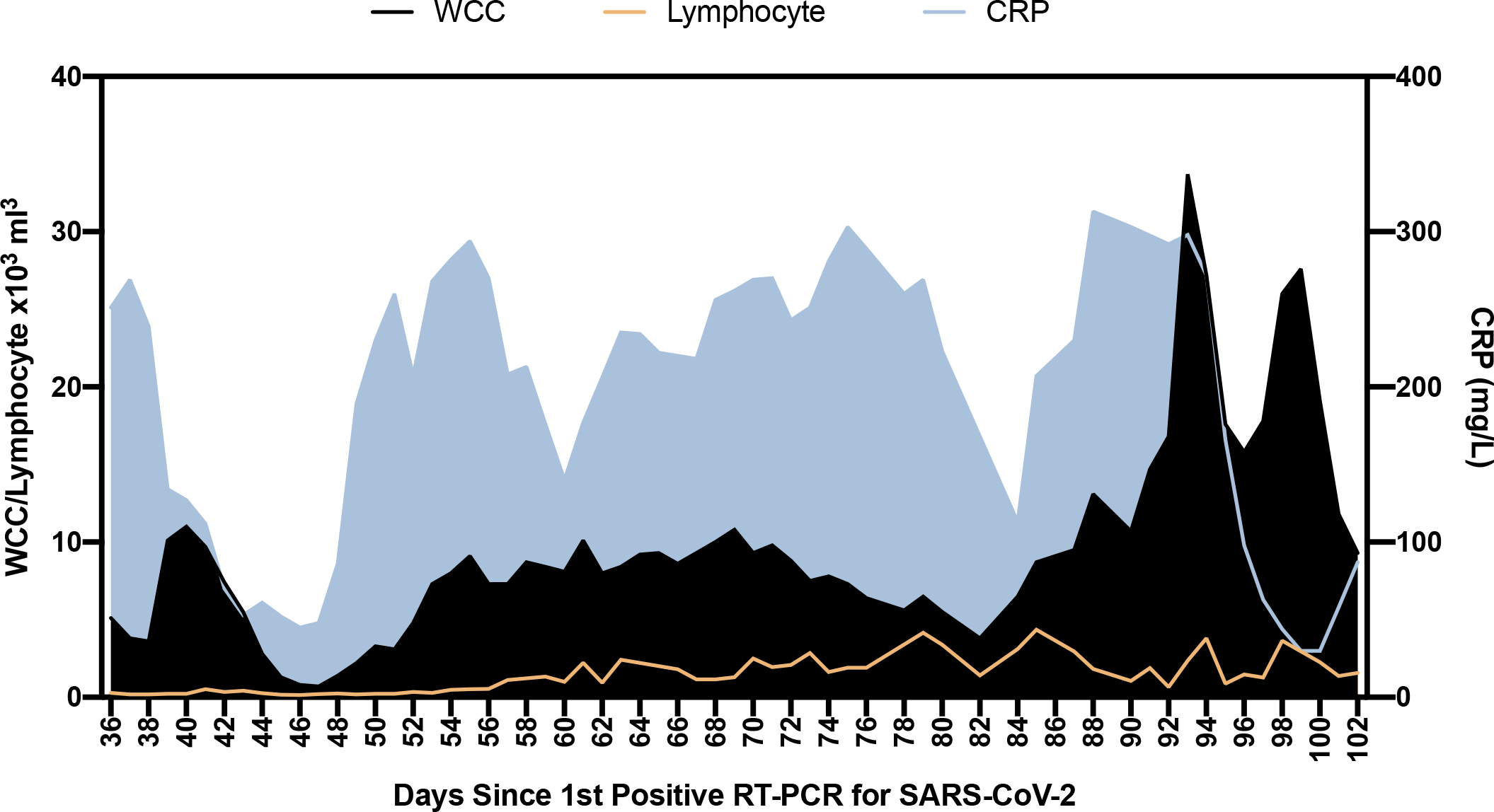
Blood parameters over time in patient case: White cell count (WCC) and lymphocyte counts are expressed as x10^3^ Cells/mm^3^. CRP: C reactive protein

**Supplementary Figure 2:**
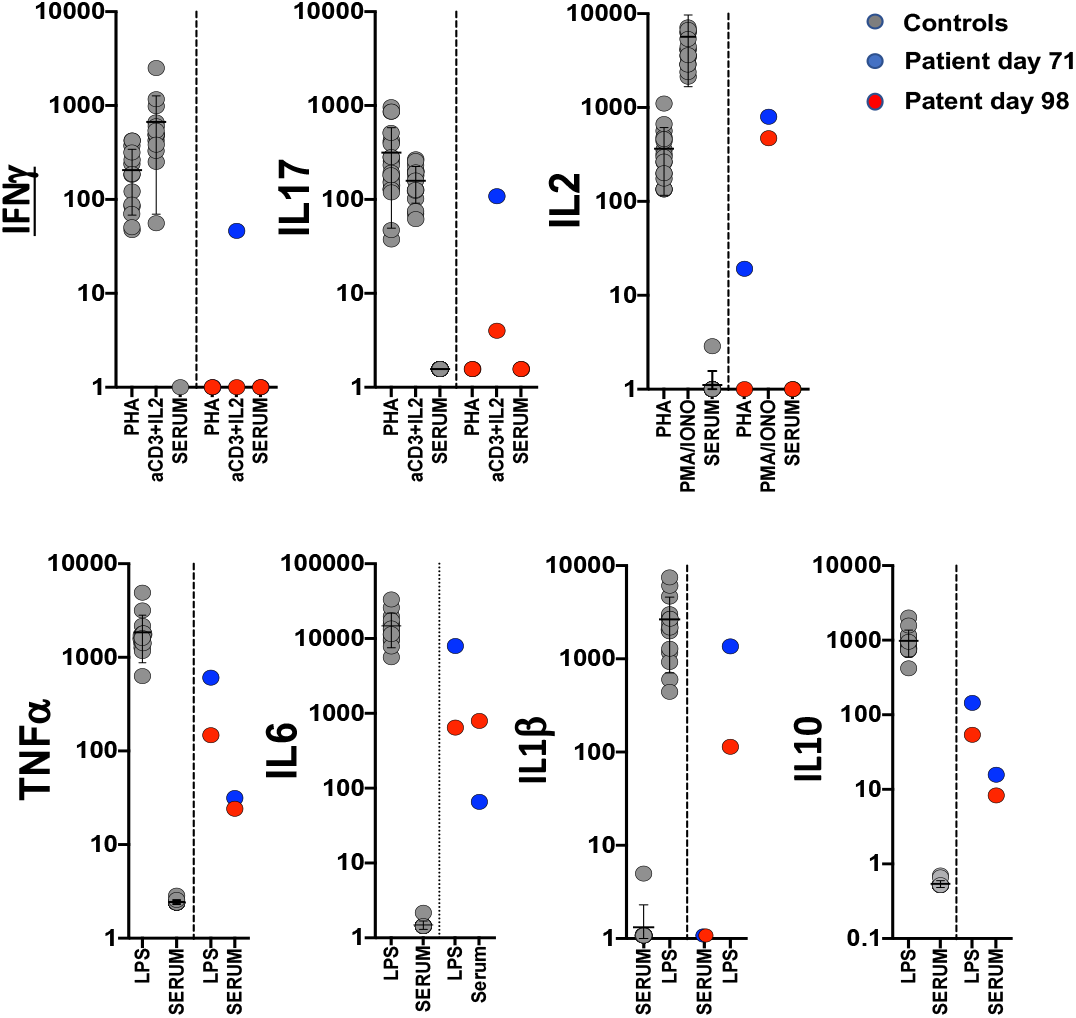
Assessment of T cell and innate function. Whole blood cytokines were measured in whole blood after 24 hours stimulation either after T-cell stimulation with PHA or anti CD3/IL2 or innate stimulation with LPS. Healthy controls are shown as grey circles (N=15), Patient at d71 and d98 is shown as blue circles or red circles respectively. Cytokine levels are shown as pg/ml. stimulation.

**Supplementary Figure 3:**
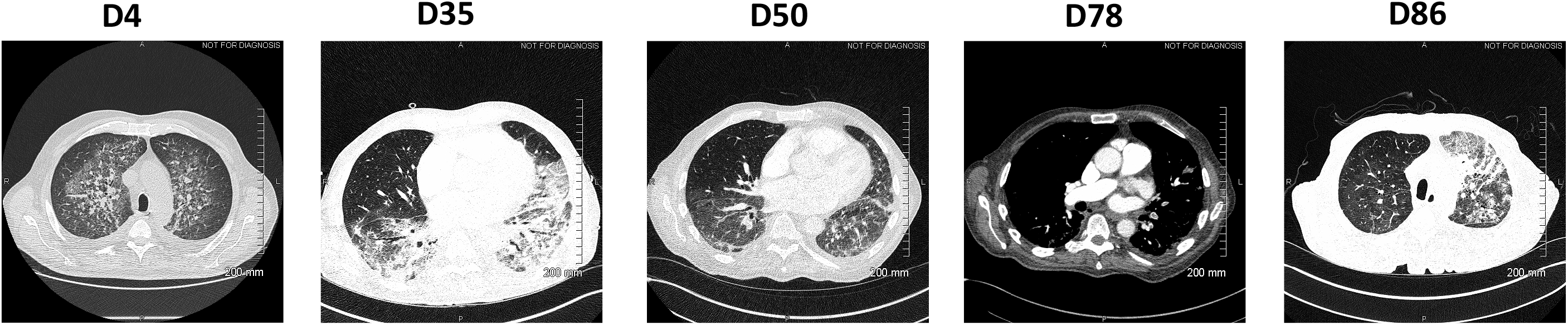
Serial CT images following detection of SARS-CoV-2. The patient initially presented with ground glass and peribroncho-vascular consolidation with associated intralobular septal thickening/reticulation and architectural distortion and interlobular septal thickening. By day 50 there is some improvement with evidence of resolving pnuemonitis, however, his condition deteriorated following the detection of bilateral pulmonary emboli, a well-recognized complication of SARS-CoV-2. Despite multiple therapeutic interventions, the patient’s condition deteriorated with worsening of inflammatory changes and chronic organizing pneumonia (COP), particularly on the left, and ongoing changes compatible with persistent SARS-CoV2 infection.

**Supplementary Figure 4:**
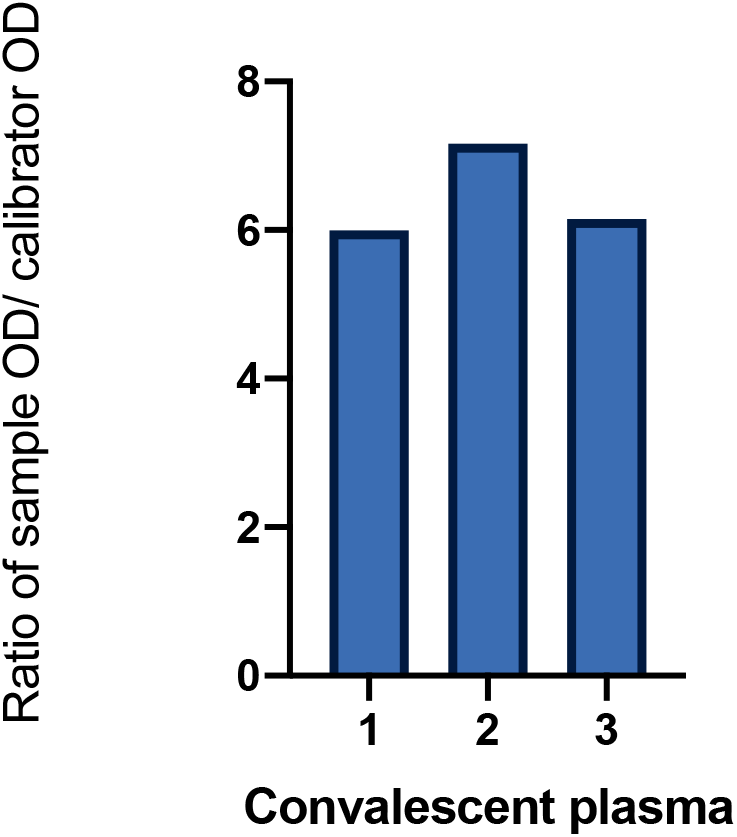
SARS-CoV-2 antibody titres in convalescent plasma. Measurement of SARS-CoV-2 specific IgG antibody titres in three units of convalescent plasma (CP) by Euroimmun assay.

**Supplementary Figure 5.**
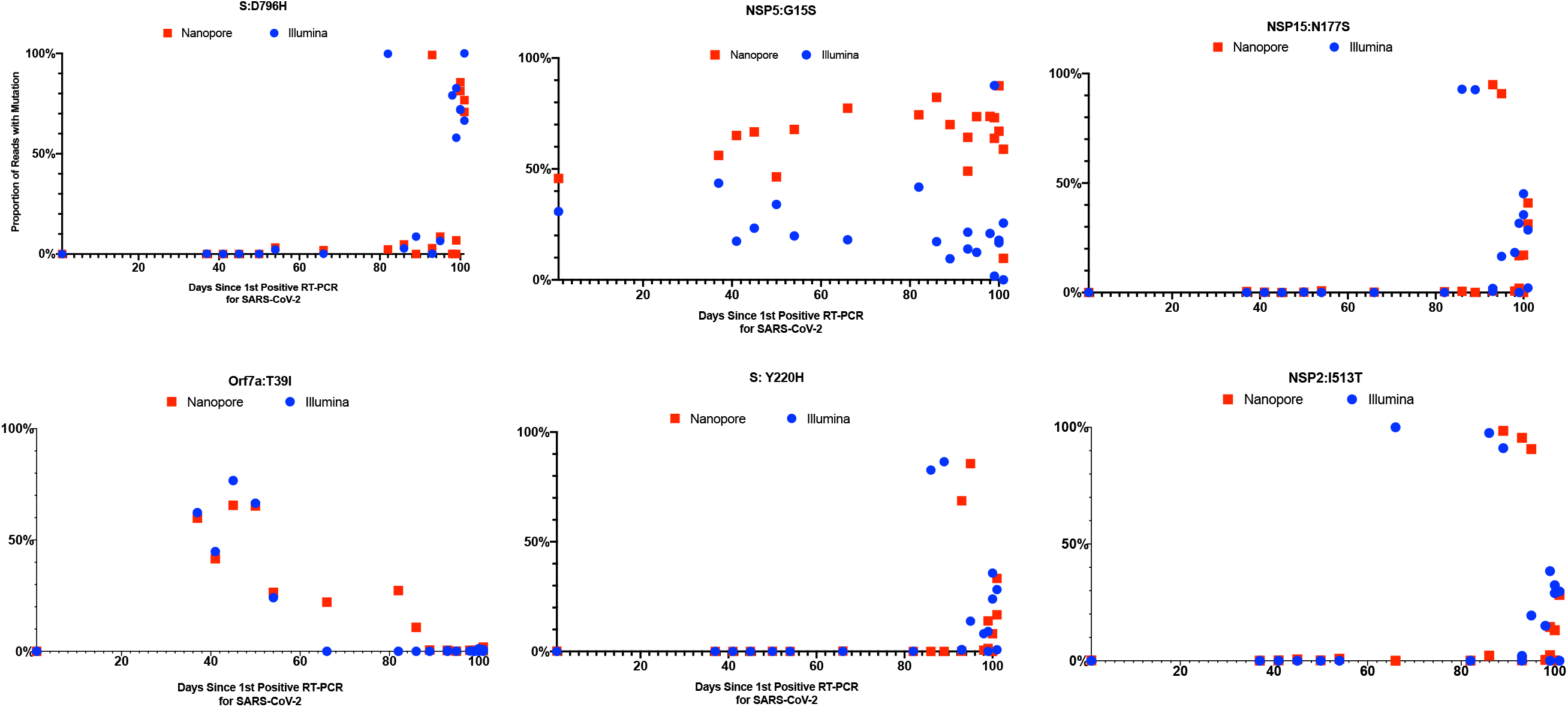
Concordance between short-read (Illumina) and long-read single molecule (Oxford Nanopore) sequencing methods for twenty samples. Points are the frequency of the variant at each timepoint; only 20 samples were sequenced using the Illumina method. Boxes represent inter-quartile ranges. Error bars are 95% CI. There was good concordance for mutations between the two methods, and no significant difference between the proportion of reads measured by both methods.

**Supplementary Figure 6:**
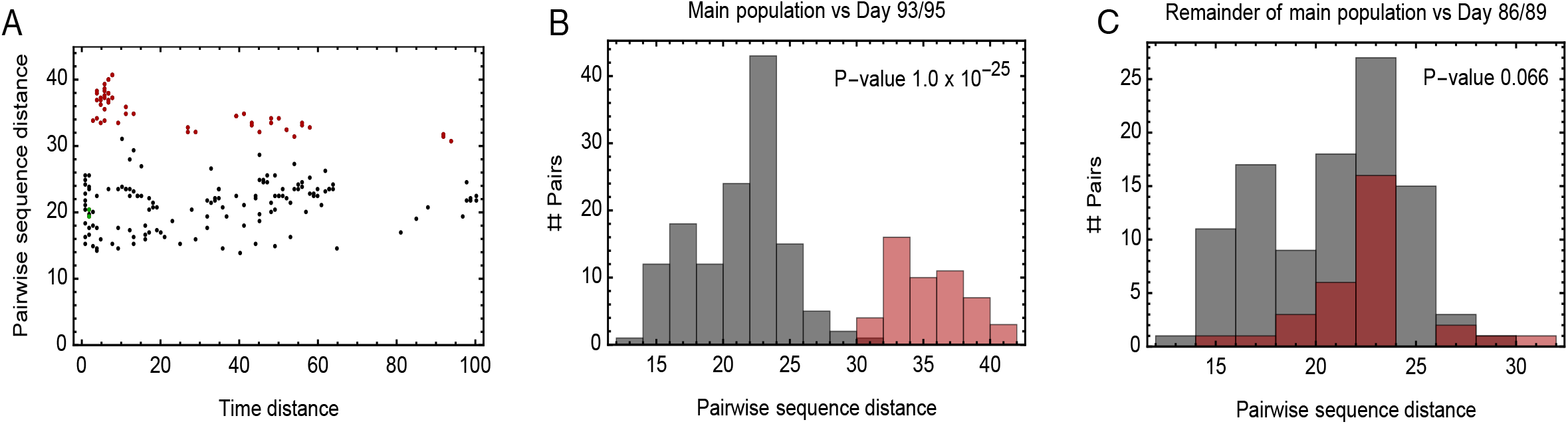
Additional evidence for within-host cladal structure. **A**. Pairwise distances between samples measured using the all-locus distance metric plotted against pairwise distances in time (measured in days) between samples being collected. Internal distances between samples in the proposed main clade are shown in black, distances between samples in the main clade and samples collected on days 93 and 95 are shown in red, and internal distances between samples collected on days 93 and 95 are shown in green. **B**. Pairwise distances between samples in the larger clade (black) and between these samples and those collected on days 93 and 95 (red). The median values of the distributions of these values are significantly different according to a Mann Whitney test. **C**. Pairwise distances between samples in the main clade, once those collected on days 86, 89, 93, 95 have been removed (black) and between these samples and those collected on days 86 and 89 (red). The median values of the distributions of these values are not significantly different at the 5% level according to a Mann Whitney test.

**Supplementary Figure 7:**
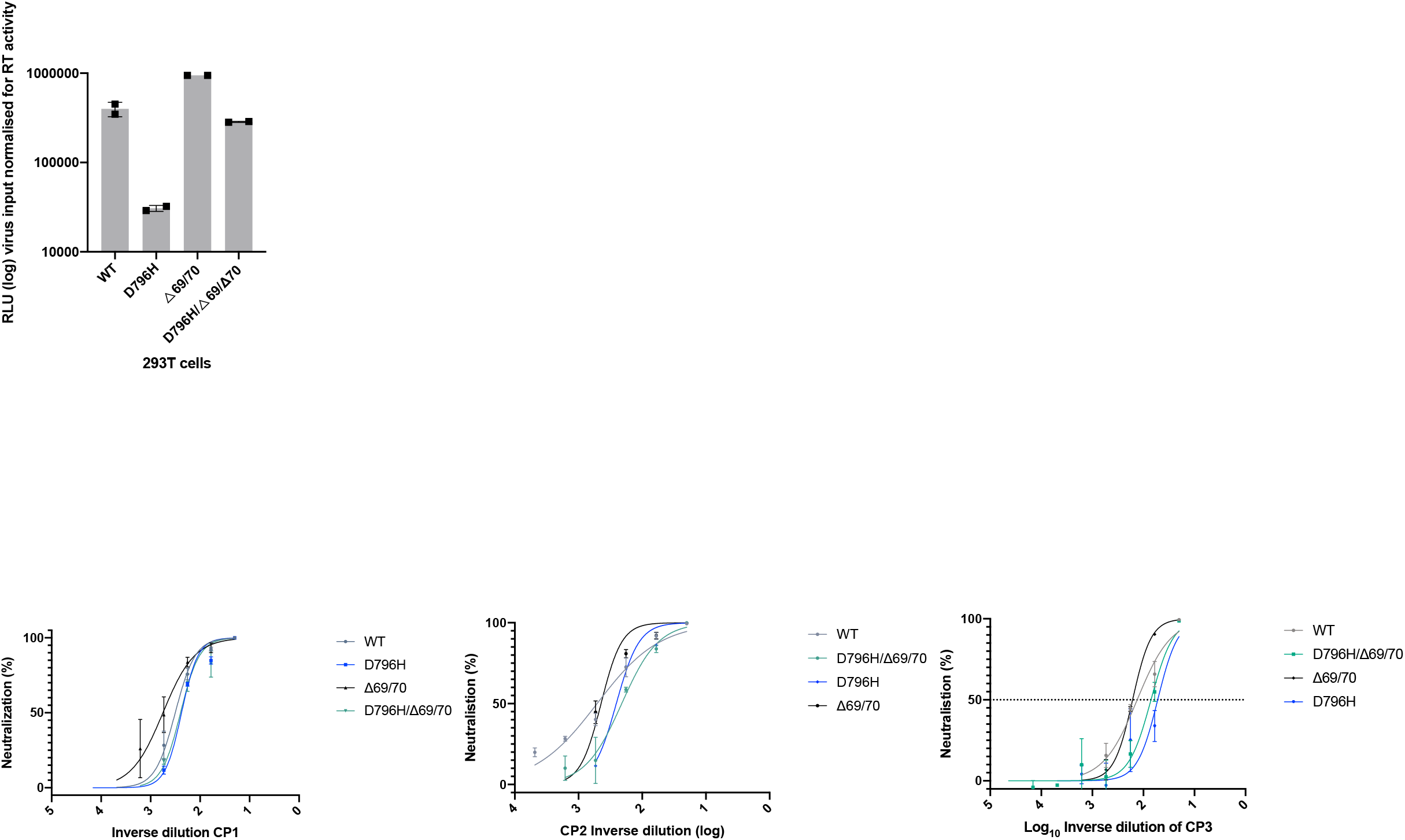
In vitro infectivity and neutralisation sensitivity of Spike pseudotyped lentiviruses. Top: infection of target 293T cells expressing TMPRSS2 and ACE2 receptors using equal amounts of virus as determined by reverse transcriptase activity. Bottom: Representative Inverse dilution plots for Spike variants against convalescent plasma units 1-3. Data points represent mean % neutralisation and error bars represent standard error of the mean

**Supplementary Figure 8.**
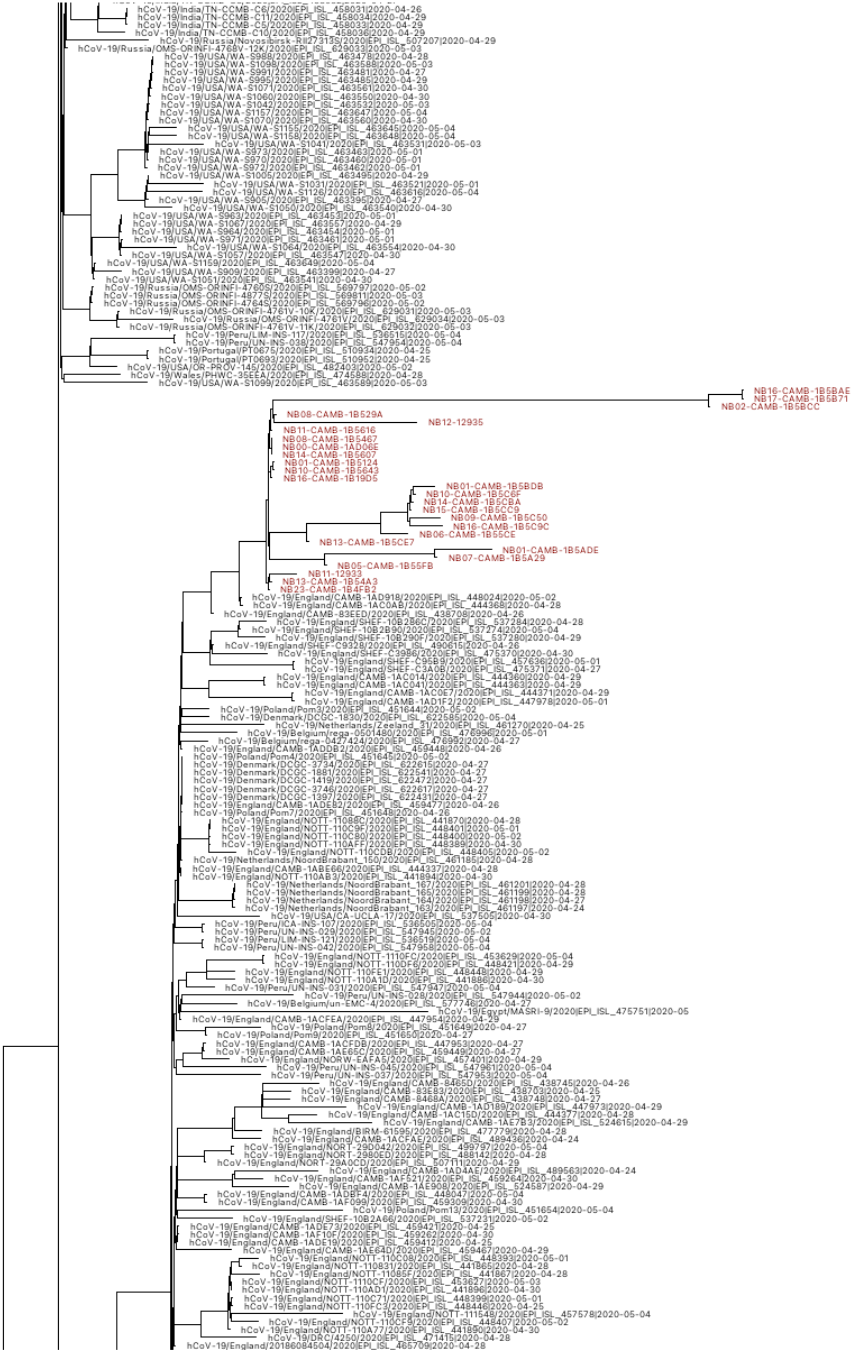
Maximum-Likelihood global phylogeny of SARS-CoV-2. To ensure that the patient case was not resultant of a superinfection event, all patient sequences were aligned with a snapshot of global SARS-CoV-2 sequences downloaded from the GISAID database between 1^st^ April and 5^th^ May (26472 sequences, only full sequences and excluding all low coverage sequences). Sequences were aligned using MAFFT and a maximum-likelihood tree inferred with IQTREE v2.1.3. All 23 sequenced from the patient case (red) formed a distinct clade, suggesting that all viral populations diversified within-host.

