## supplementary information for "Neutralising antibodies in Spike mediated SARS-CoV-2 adaptation"

| Component | Reference range | Day 49 | Day 89 |
|---|---|---|---|
| Lymphocyte count | 1.00 – 2.80 X $10^9$/l | 0.16 | 1.97 |
| CD3 % | % | 49 | 22 |
| CD3 total | 0.70 – 2.10 X $10^9$/l | 0.08 | 0.43 |
| CD4 % | % | 29 | 11 |
| CD4 total | 0.30 – 1.40 X $10^9$/l | 0.05 | 0.22 |
| CD8 % | % | 21 | 10 |
| CD8 total | 0.20 – 0.90 X $10^9$/l | 0.03 | 0.2 |
| CD19 % | % | 21 | 58 |
| CD19 total | 0.10 – 0.50 X $10^9$/l | 0.03 | 1.14 |
| CD56 % | % | 23 | 19 |
| CD56 total | 0.09 – 0.60 X $10^9$/l | 0.04 | 0.37 |
| IgG | 6.00 – 16.00 g/l | <1.40 | 1.9 |
| IgA | 0.80 – 4.00 g/l | <0.30 | <0.30 |
| IgM | 0.50 – 2.00 g/l | 1.27 | 1.92 |
| Sars-CoV-2 Total antibody | (Siemens) | Negative | Positive |

**Supplemental Table 1**: Immunological parameters of patient pre- and post- receipt of convalescent plasma (CP) demonstrating profound lymphopenia with reduction in total serum IgG and IgA. The patient continued to have normal serum IgM which either reflects his underlying lymphoproliferative state or ongoing infection. Following receipt of CP the presence of SARS-CoV-2 antibodies is evident.

**Supplementary Table 2. Characteristics of genomes used in the phylogenetic analysis in Figure 2B.** Pangolin lineages were identified using Pangolin COVID-19 Lineage Assigner v2.0.8 (https://pangolin.cog-uk.io/).. CP- convalescent plasma; mAb – monoclonal antibody.

| Patient | Underlying Clinical Diagnosis | Pangolin Lineage | Treatments |
|---|---|---|---|
| **Global GSAID Average** | - | - | NA |
| **Choi et al** | Antiphospholipid syndrome | B.1 | Dual mAb, 3 x remdesivir |
| **Avanzato et al** | Chronic lymphocytic leukemia and HLA | A.1 | 2x CP units |
| **Control patient 1 (purple)** | X-linked Agammaglobulinemia | B.2.6 | 2 x remdesevir, CP after virus clearance |
| **Control Patient 2 (blue)** | Chronic renal impairment | B.1.1 | None |
| **Control Patient 3 (red)** | Heart disease | B.1.1.35 | None |
| **Study patient** | Marginal B Cell Lymphoma | B.1.1.35/1.1.1 | 3x CP units, 3 x remdesivir |

| Timepoint (Day) | sequence | CT value | Sample type | Nanopore | Illumina | SGA |
|---|---|---|---|---|---|---|
| 1 | NB16_CAMB-1B19D5 | 25 | NT | Y | Y | Y |
| 37 | NB23_CAMB-1B4FB2 | NA | NT | Y | Y | Y |
| 41 | NB01_CAMB-1B5124 | 23 | NT | Y | Y | - |
| 45 | NB08_CAMB-1B529A | 29 | NT | Y | Y | - |
| 50 | NB13_CAMB-1B54A3 | NA | NT | Y | Y | - |
| 54 | NB10_CAMB-1B5643 | 22 | NT | Y | - | - |
| 55 | NB08_CAMB-1B5467 | NA | NT | Y | Y | Y |
| 56 | NB11_CAMB-1B5616 | 23 | NT | Y | - | - |
| 57 | NB14_CAMB-1B5607 | 24 | NT | Y | - | - |
| 66 | NB05_CAMB-1B55FB | 26 | NT | Y | Y | - |
| 82 | NB06_CAMB-1B55CE | 23 | NT | Y | Y | - |
| 89 | NB01_CAMB-1B5ADE | NA | NT | Y | Y | - |
| 93 | NB16_CAMB-1B5BAE | 21 | NT | Y | Y | - |
| 93 | NB17_CAMB-1B5B71 | 19 | ETA | Y | Y | - |
| 95 | NB02_CAMB-1B5BCC | 16 | ETA | Y | Y | - |
| 98 | NB01_CAMB-1B5BDB | 25 | NT | Y | Y | Y |
| 99 | NB09_CAMB-1B5C50 | 30 | NT | Y | Y | - |
| 99 | NB10_CAMB-1B5C6F | 18 | ETA | Y | Y | - |
| 100 | NB13_CAMB-1B5CE7 | NA | NT | Y | Y | - |
| 100 | NB14_CAMB-1B5CBA | 19 | ETA | Y | Y | - |
| 101 | NB15_CAMB-1B5CC9 | 24 | ETA | Y | Y | - |
| 101 | NB16_CAMB-1B5C9C | 30 | NT | Y | Y | - |

**Supplementary table 3 : Samples and sequencing methods**. Timepoint indicates the day since 1$^{st}$ positive qPCR for SARS-COV-2 that the sample was taken. CT values are reported where available. Y- Yes, - Not done, NA not available

**Supplementary Table 4. Prevalence of selected Spike glycoprotein mutations at sequential time points** and sequencing depth (number of reads covering the amino acid position) measured by both short-read (Illumina) and long-read (Oxford Nanopore) methods. There was low coverage of S:P330S and S:D796H in the final three timepoints, measured by both short- and long-read methods.

| | | | | Illumina | Nanopore | Illumina | Nanopore | Illumina | Nanopore | Illumina | Nanopore | Illumina | Nanopore | Illumina | Nanopore | Illumina | Nanopore |
|---|---|---|---|---|---|---|---|---|---|---|---|---|---|---|---|---|---|
| | | | Day | 1 | 1 | 37 | 37 | 41 | 41 | 45 | 45 | 50 | 50 | 54 | 54 | 66 | 66 |
| Locus | Position | From | To | | | | | | | | | | | | | | |
| 2343 | NSP2 | T | C | 0.0% | 0.2% | 0.0% | 0.1% | 0.0% | 0.2% | 0.0% | 0.6% | 0.0% | 0.2% | 0.0% | 0.9% | 100.0% | 0.0% |
| 3057 | NSP3 | A | G | 0.0% | 0.0% | 0.0% | 0.0% | 0.0% | 0.0% | 0.0% | 0.0% | 0.0% | 0.0% | 0.2% | 0.0% | 0.0% | 0.0% |
| 5393 | NSP3 | T | C | 0.0% | 1.4% | 0.2% | 0.7% | 0.0% | 1.1% | 0.0% | 0.0% | 0.2% | 1.0% | 0.0% | 0.9% | 0.0% | 1.0% |
| 5406 | NSP3 | T | G | 0.0% | 0.2% | 0.0% | 0.0% | 0.0% | 0.0% | 0.0% | 0.0% | 0.2% | 0.0% | 0.0% | 0.0% | 0.1% | 0.0% |
| 5425 | NSP3 | G | T | 0.0% | 0.0% | 0.0% | 0.1% | 0.0% | 0.1% | 0.0% | 0.0% | 0.0% | 0.1% | 0.0% | 0.1% | 0.0% | 0.0% |
| 8389 | NSP3 | C | T | 0.0% | 0.9% | 0.0% | 0.4% | 0.1% | 0.5% | 0.0% | 0.0% | 0.0% | 0.9% | 0.0% | 0.4% | 0.0% | 0.5% |
| 9130 | NSP4 | G | T | 0.0% | 0.1% | 0.0% | 0.1% | 0.0% | 0.2% | 0.0% | 0.0% | 0.0% | 0.2% | 0.0% | 0.3% | 0.0% | 0.4% |
| 10097 | NSP5 | A | G | 30.8% | 45.7% | 43.6% | 56.1% | 17.4% | 65.1% | 23.3% | 66.7% | 34.0% | 46.4% | 19.8% | 67.8% | 18.1% | 77.4% |
| 10700 | NSP5 | G | A | 0.0% | 0.5% | 0.0% | 0.7% | 0.1% | 0.7% | 0.0% | 0.0% | 0.0% | 0.7% | 0.1% | 0.9% | 0.0% | 0.8% |
| 11770 | NSP6 | A | G | 0.0% | 0.7% | 0.0% | 0.4% | 0.1% | 0.2% | 0.0% | 2.2% | 0.1% | 0.7% | 0.0% | 0.2% | 0.0% | 0.4% |
| 12043 | NSP7 | C | T | 0.0% | 0.7% | 0.1% | 0.5% | 0.0% | 0.7% | 0.0% | 0.0% | 0.1% | 0.5% | 0.0% | 0.9% | 0.0% | 0.7% |
| 13527 | NSP12b | T | C | 0.0% | 0.0% | 0.0% | 0.1% | 0.0% | 0.0% | 0.0% | 0.0% | 0.0% | 0.1% | 0.0% | 0.1% | 0.0% | 0.1% |
| 14776 | NSP12b | G | T | 0.0% | 0.5% | 0.0% | 0.3% | 0.0% | 0.2% | 0.1% | 0.0% | 0.2% | 0.1% | 0.0% | 0.1% | 0.1% | 0.1% |
| 16733 | NSP13 | C | T | 0.1% | 0.2% | 2.6% | 1.7% | 20.9% | 13.2% | 37.0% | 29.8% | 0.0% | 0.3% | 3.6% | 1.3% | 0.0% | 2.0% |
| 17320 | NSP13 | G | T | 0.1% | 0.3% | 0.3% | 0.1% | 0.1% | 0.2% | 0.0% | 0.7% | 0.0% | 0.1% | 0.0% | 0.0% | 0.0% | 0.1% |
| 17703 | NSP13 | C | T | 0.0% | 0.6% | 0.1% | 0.8% | 0.0% | 1.3% | 0.0% | 1.9% | 0.0% | 1.5% | 0.0% | 1.0% | 0.0% | 1.0% |
| 18488 | NSP14a | T | C | 0.0% | 0.3% | 0.0% | 0.2% | 0.1% | 0.4% | 0.0% | 0.0% | 0.0% | 0.3% | 0.0% | 0.2% | 0.0% | 0.3% |
| 19388 | NSP14a | G | A | 0.0% | 0.0% | 0.0% | 0.0% | 0.0% | 0.2% | 0.0% | 3.1% | 0.0% | 0.4% | 0.0% | 0.2% | 0.0% | 0.5% |
| 20150 | NSP15 | A | G | 0.0% | 0.0% | 0.0% | 0.5% | 0.0% | 0.2% | 0.0% | 0.0% | 0.2% | 0.2% | 0.0% | 0.8% | 0.0% | 0.2% |
| 21600 | S | G | T | 0.1% | 0.0% | 0.0% | 0.0% | 0.0% | 0.0% | 0.0% | 0.0% | 0.1% | 0.0% | 0.0% | 0.0% | 0.1% | 0.0% |
| 21635 | S | C | A | 0.0% | 0.0% | 0.0% | 0.0% | 0.0% | 0.7% | 0.0% | 0.0% | 0.0% | 1.4% | 0.0% | 0.0% | 0.2% | 0.8% |
| 21753 | S | T | G | 0.0% | 0.4% | 0.0% | 0.8% | 0.0% | 0.0% | 0.0% | 0.7% | 0.0% | 0.0% | 0.0% | 0.4% | 0.0% | 0.0% |
| 21768 | S | | - | 0.0% | 8.4% | 0.6% | 0.1% | 0.0% | 0.0% | 0.0% | 0.0% | 0.0% | 0.0% | 0.0% | 0.1% | 0.0% | 4.4% |

| | | | | | | | | | | | | | | | | | |
|---|---|---|---|---|---|---|---|---|---|---|---|---|---|---|---|---|---|
| 22088 | S | C | T | 0.1% | 1.0% | 0.0% | 0.3% | 0.0% | 0.4% | 0.0% | 0.0% | 0.0% | 0.6% | 0.5% | 0.7% | 0.0% | 0.9% |
| 22160 | S | T | C | 0.0% | 0.2% | 0.0% | 0.0% | 0.0% | 0.0% | 0.0% | 0.0% | 0.0% | 0.1% | 0.0% | 0.0% | 0.0% | 0.2% |
| 22281 | S | C | T | 0.1% | 0.2% | 0.0% | 0.1% | 0.0% | 0.1% | 0.0% | 0.0% | 0.0% | 0.1% | 0.0% | 0.4% | 0.0% | 0.1% |
| 22551 | S | C | T | 0.0% | 1.1% | 0.0% | 0.8% | 0.0% | 0.0% | 0.0% | 0.7% | 0.0% | 0.0% | 0.1% | 1.6% | 0.0% | 9.0% |
| 23398 | S | T | C | 0.0% | 0.3% | 0.0% | 0.2% | 0.2% | 0.1% | 0.0% | 0.7% | 0.0% | 0.0% | 0.1% | 0.1% | 0.1% | 0.2% |
| 23948 | S | G | C | 0.0% | 0.0% | 0.2% | 0.0% | 0.0% | 0.0% | 0.1% | 0.0% | 0.1% | 0.0% | 2.2% | 3.2% | 0.2% | 1.9% |
| 24257 | S | G | T | 0.0% | 0.0% | 0.0% | 0.0% | 2.1% | 2.3% | 0.1% | 2.7% | 27.0% | 25.7% | 3.3% | 0.6% | 0.0% | 2.6% |
| 25537 | E | G | A | 0.0% | 0.3% | 2.6% | 2.8% | 16.5% | 15.4% | 20.3% | 20.8% | 0.1% | 0.2% | 2.8% | 1.0% | 0.0% | 2.4% |
| 26333 | E | C | T | 0.0% | 0.6% | 0.0% | 0.1% | 0.0% | 0.4% | 0.0% | 0.0% | 0.0% | 0.2% | 0.2% | 0.5% | 0.0% | 0.3% |
| 26634 | M | G | T | 0.1% | 0.4% | 0.0% | 0.2% | 0.0% | 0.2% | 0.0% | 0.8% | 0.0% | 0.3% | 0.0% | 0.2% | 0.0% | 0.2% |
| 26647 | M | G | A | 0.0% | 0.0% | 0.0% | 0.2% | 0.0% | 0.2% | 0.0% | 0.9% | 0.0% | 0.1% | 0.0% | 0.0% | 0.0% | 0.0% |
| 27087 | M | G | A | 0.0% | 0.6% | 0.0% | 0.3% | 0.1% | 0.1% | 0.0% | 0.0% | 0.0% | 0.1% | 0.0% | 0.2% | 0.0% | 0.1% |
| 27408 | ORF7a | T | C | 0.0% | 0.0% | 0.1% | 0.0% | 0.0% | 0.2% | 0.0% | 0.0% | 0.0% | 0.0% | 0.0% | 0.0% | 0.1% | 0.1% |
| 27459 | ORF7a | G | T | 0.1% | 0.4% | 0.1% | 0.0% | 0.1% | 0.1% | 0.0% | 1.4% | 0.1% | 0.1% | 0.0% | 0.2% | 0.0% | 0.4% |
| 27509 | ORF7a | C | T | 0.0% | 0.0% | 62.3% | 59.8% | 44.8% | 41.6% | 76.7% | 65.6% | 66.5% | 65.3% | 24.1% | 26.5% | 0.0% | 22.1% |
| 27618 | ORF7a | T | C | 0.2% | 0.0% | 0.0% | 0.0% | 0.0% | 0.0% | 0.0% | 0.0% | 0.0% | 0.4% | 0.0% | 0.0% | 0.0% | 0.8% |
| 28209 | ORF7b | G | C | 0.0% | 0.2% | 0.0% | 0.0% | 0.1% | 0.1% | 0.0% | 0.4% | 0.0% | 0.0% | 0.0% | 0.0% | 0.0% | 0.0% |
| 28356 | N | A | G | 0.0% | 0.2% | 0.3% | 0.6% | 1.9% | 3.0% | 0.0% | 0.7% | 51.0% | 48.1% | 0.5% | 0.5% | 0.0% | 0.5% |
| 28748 | N | C | T | 0.0% | 0.4% | 0.0% | 0.1% | 0.1% | 0.3% | 0.0% | 0.0% | 0.0% | 0.3% | 0.2% | 0.3% | 0.0% | 0.2% |
| 29825 | 3'UTR | G | T | 0.0% | 1.3% | 0.2% | 0.3% | 0.0% | 0.6% | 0.0% | 0.0% | 0.2% | 0.4% | 0.0% | 0.2% | 0.0% | 0.7% |

| | | | | Illumina | Nanopore | Illumina | Nanopore | Illumina | Nanopore | Illumina | Nanopore | Illumina | Nanopore | Illumina | Nanopore | Illumina | Nanopore |
|---|---|---|---|---|---|---|---|---|---|---|---|---|---|---|---|---|---|
| | | | Day | 82 | 82 | 86 | 86 | 89 | 89 | 93 | 93 | 93 | 93 | 95 | 95 | 98 | 98 |
| Locus | Position | From | To | | | | | | | | | | | | | | |
| 2343 | NSP2 | T | C | 0.0% | 0.1% | 97.6% | 2.2% | 91.1% | 98.5% | 0.1% | 0.2% | 2.1% | 95.5% | 19.4% | 90.7% | 15.0% | 0.4% |
| 3057 | NSP3 | A | G | 0.0% | 0.1% | 0.0% | 0.0% | 0.0% | 0.1% | 0.1% | 0.0% | 0.0% | 0.0% | 0.0% | 0.0% | 0.7% | 0.1% |
| 5393 | NSP3 | T | C | 0.2% | 1.3% | 0.4% | 0.9% | 0.0% | 1.3% | 0.2% | 1.2% | 0.0% | 1.3% | 7.4% | 1.2% | 78.8% | 1.2% |
| 5406 | NSP3 | T | G | 0.0% | 0.0% | 29.3% | 0.1% | 66.5% | 0.0% | 0.3% | 0.1% | 2.3% | 15.8% | 17.4% | 56.2% | 10.9% | 0.3% |
| 5425 | NSP3 | G | T | 99.9% | 0.3% | 0.9% | 0.0% | 9.0% | 0.0% | 0.0% | 97.4% | 0.6% | 0.5% | 7.4% | 7.9% | 82.6% | 0.2% |
| 8389 | NSP3 | C | T | 0.0% | 0.7% | 0.0% | 0.3% | 0.0% | 0.3% | 20.6% | 0.5% | 19.6% | 0.7% | 22.2% | 0.4% | 0.1% | 21.9% |
| 9130 | NSP4 | G | T | 0.0% | 0.4% | 0.0% | 0.2% | 0.0% | 0.3% | 0.1% | 0.3% | 0.0% | 0.3% | 0.0% | 0.3% | 12.6% | 0.4% |
| 10097 | NSP5 | A | G | 41.8% | 74.4% | 17.2% | 82.3% | 9.5% | 70.0% | 21.5% | 49.0% | 13.9% | 64.3% | 12.4% | 73.6% | 20.9% | 73.7% |
| 10700 | NSP5 | G | A | 0.0% | 0.4% | 0.0% | 0.6% | 0.3% | 0.8% | 99.5% | 0.6% | 96.8% | 0.6% | 76.9% | 0.5% | 0.1% | 98.8% |
| 11770 | NSP6 | A | G | 0.1% | 0.5% | 0.3% | 0.4% | 0.1% | 0.4% | 97.9% | 0.3% | 90.4% | 0.4% | 73.1% | 0.3% | 0.1% | 96.8% |
| 12043 | NSP7 | C | T | 0.0% | 1.1% | 20.1% | 0.4% | 51.1% | 0.6% | 0.2% | 0.7% | 3.3% | 20.7% | 13.0% | 53.3% | 13.3% | 0.6% |
| 13527 | NSP12b | T | C | 0.0% | 0.2% | 0.3% | 0.1% | 0.4% | 0.0% | 57.3% | 0.0% | 52.1% | 0.1% | 34.8% | 0.1% | 0.0% | 54.6% |
| 14776 | NSP12b | G | T | 0.1% | 0.2% | 0.2% | 0.1% | 0.0% | 0.1% | 54.1% | 0.3% | 50.7% | 0.6% | 34.6% | 0.2% | 0.0% | 57.7% |
| 16733 | NSP13 | C | T | 0.0% | 5.0% | 0.0% | 2.6% | 0.0% | 0.2% | 0.0% | 0.1% | 0.0% | 0.2% | 0.0% | 0.2% | 0.4% | 0.1% |
| 17320 | NSP13 | G | T | 0.0% | 0.1% | 0.5% | 0.0% | 0.0% | 0.1% | 0.0% | 0.1% | 0.0% | 0.5% | 0.0% | 0.2% | 1.0% | 0.1% |
| 17703 | NSP13 | C | T | 0.0% | 1.4% | 0.0% | 1.5% | 0.0% | 0.8% | 15.5% | 1.4% | 9.0% | 1.0% | 17.4% | 1.1% | 0.0% | 16.1% |
| 18488 | NSP14a | T | C | 0.0% | 0.2% | 0.0% | 0.2% | 0.0% | 0.4% | 16.2% | 0.2% | 8.5% | 0.3% | 18.3% | 0.3% | 0.0% | 15.7% |
| 19388 | NSP14a | G | A | 0.0% | 0.2% | 0.0% | 0.2% | 0.0% | 0.2% | 17.8% | 0.5% | 11.9% | 0.7% | 20.9% | 0.0% | 0.0% | 17.4% |
| 20150 | NSP15 | A | G | 0.0% | 0.4% | 92.8% | 0.6% | 92.6% | 0.1% | 0.6% | 0.7% | 1.9% | 94.9% | 16.5% | 90.8% | 18.3% | 0.7% |
| 21600 | S | G | T | 0.0% | 0.0% | 0.0% | 0.0% | 0.3% | 0.0% | 99.2% | 0.0% | 97.1% | 0.0% | 76.3% | 0.0% | 0.7% | 99.1% |
| 21635 | S | C | A | 0.0% | 0.5% | 0.0% | 0.0% | 0.2% | 1.3% | 27.9% | 0.8% | 14.6% | 0.4% | 23.5% | 0.0% | 0.0% | 26.8% |
| 21753 | S | T | G | 0.0% | 0.7% | 0.0% | 0.8% | 0.0% | 0.5% | 96.5% | 80.2% | 0.0% | 0.0% | 62.7% | 60.3% | 0.0% | 0.3% |
| 21768 | S | | - | 99.0% | 93.0% | 0.0% | 6.9% | 6.0% | 1.0% | 0.0% | 9.2% | 0.0% | 0.0% | 12.0% | 10.4% | 82.6% | 93.5% |
| 22088 | S | C | T | 0.1% | 0.9% | 0.0% | 0.9% | 0.0% | 1.1% | 15.6% | 1.1% | 6.4% | 2.8% | 16.8% | 1.2% | 0.0% | 6.8% |
| 22160 | S | T | C | 0.1% | 0.0% | 82.7% | 0.0% | 86.5% | 0.0% | 0.6% | 0.1% | 0.9% | 68.7% | 13.8% | 85.6% | 8.1% | 0.6% |
| 22281 | S | C | T | 0.1% | 0.3% | 82.7% | 0.2% | 84.5% | 0.1% | 0.4% | 0.2% | 1.8% | 54.0% | 11.1% | 73.3% | 6.3% | 0.3% |
| 22551 | S | C | T | 0.0% | 0.9% | 0.0% | 0.9% | 0.0% | 0.9% | 96.9% | 92.8% | 0.0% | 0.0% | 78.3% | 73.3% | 0.0% | 1.3% |

| | | | | | | | | | | | | | | | | | |
|---|---|---|---|---|---|---|---|---|---|---|---|---|---|---|---|---|---|
| 23398 | S | T | C | 0.1% | 0.0% | 24.9% | 0.0% | 65.9% | 0.1% | 0.4% | 0.1% | 2.2% | 22.1% | 16.2% | 62.1% | 12.7% | 0.4% |
| 23948 | S | G | C | 99.8% | 2.3% | 2.8% | 4.8% | 8.7% | 0.0% | 0.2% | 99.2% | 0.2% | 2.9% | 6.5% | 8.7% | 79.1% | 0.1% |
| 24257 | S | G | T | 0.0% | 0.4% | 0.0% | 0.3% | 0.0% | 0.4% | 0.0% | 0.3% | 0.0% | 0.1% | 0.0% | 0.2% | 0.0% | 0.0% |
| 25537 | E | G | A | 0.0% | 3.5% | 0.0% | 1.4% | 0.0% | 0.1% | 0.0% | 0.1% | 0.0% | 0.1% | 0.0% | 0.2% | 0.0% | 0.1% |
| 26333 | E | C | T | 0.0% | 0.5% | 0.0% | 0.2% | 0.2% | 0.2% | 99.3% | 0.5% | 97.6% | 0.3% | 77.9% | 0.1% | 0.0% | 97.8% |
| 26634 | M | G | T | 0.1% | 0.2% | 0.1% | 0.2% | 0.0% | 0.1% | 43.0% | 0.3% | 31.8% | 0.2% | 10.6% | 0.2% | 0.0% | 40.2% |
| 26647 | M | G | A | 0.0% | 0.1% | 0.0% | 0.1% | 0.0% | 0.1% | 22.5% | 0.1% | 27.6% | 0.1% | 6.5% | 0.1% | 0.0% | 18.5% |
| 27087 | M | G | A | 0.1% | 0.3% | 0.0% | 0.2% | 0.0% | 0.1% | 16.4% | 0.3% | 10.7% | 0.3% | 18.2% | 0.2% | 0.0% | 18.6% |
| 27408 | ORF7a | T | C | 0.0% | 0.1% | 28.2% | 0.0% | 64.1% | 0.0% | 0.3% | 0.1% | 2.5% | 23.3% | 14.0% | 49.5% | 10.0% | 0.3% |
| 27459 | ORF7a | G | T | 0.2% | 0.3% | 0.0% | 0.7% | 2.6% | 0.1% | 0.1% | 0.3% | 0.1% | 0.0% | 4.1% | 3.5% | 66.9% | 0.2% |
| 27509 | ORF7a | C | T | 0.0% | 27.3% | 0.0% | 10.8% | 0.0% | 0.6% | 0.0% | 0.5% | 0.4% | 0.4% | 0.1% | 0.1% | 0.2% | 0.5% |
| 27618 | ORF7a | T | C | 0.1% | 0.0% | 0.0% | 0.0% | 1.4% | 0.0% | 99.5% | 0.3% | 97.4% | 0.0% | 85.6% | 0.0% | 0.8% | 97.5% |
| 28209 | ORF7b | G | C | 0.0% | 0.0% | 0.0% | 0.0% | 0.7% | 0.0% | 0.2% | 0.0% | 1.5% | 0.0% | 12.4% | 0.6% | 3.1% | 0.3% |
| 28356 | N | A | G | 99.9% | 0.6% | 2.3% | 0.6% | 6.1% | 0.6% | 0.2% | 98.3% | 0.2% | 2.7% | 5.9% | 6.6% | 78.6% | 0.6% |
| 28748 | N | C | T | 0.0% | 0.4% | 2.2% | 0.2% | 0.0% | 0.3% | 0.1% | 0.3% | 1.7% | 2.6% | 12.9% | 0.1% | 2.8% | 0.4% |
| 29825 | 3'UTR | G | T | 99.8% | 0.5% | 4.7% | 0.4% | 8.7% | 0.1% | 0.3% | 98.5% | 0.5% | 3.0% | 6.4% | 9.4% | 80.5% | 0.7% |

| | | | | Illumina | Nanopore | Illumina | Nanopore | Illumina | Nanopore | Illumina | Nanopore | Illumina | Nanopore | Illumina | Nanopore |
|---|---|---|---|---|---|---|---|---|---|---|---|---|---|---|---|
| | | | Day | 99 | 99 | 99 | 99 | 100 | 100 | 100 | 100 | 101 | 101 | 101 | 101 |
| Locus | Position | From | To | | | | | | | | | | | | |
| 2343 | NSP2 | T | C | 18.2% | 81.4% | 5.4% | 67.1% | 0.0% | 0.4% | 2.8% | 11.9% | 7.8% | 3.1% | 0.0% | 0.6% |
| 3057 | NSP3 | A | G | 0.0% | 2.4% | 38.4% | 14.5% | 29.0% | 13.1% | 32.4% | 0.2% | 29.7% | 28.6% | 0.0% | 28.2% |
| 5393 | NSP3 | T | C | 14.4% | 0.0% | 0.0% | 0.0% | 10.4% | 0.1% | 0.0% | 12.3% | 0.0% | 0.0% | 0.0% | 9.5% |
| 5406 | NSP3 | T | G | 46.4% | 1.4% | 65.3% | 7.4% | 30.8% | 77.0% | 65.3% | 39.3% | 65.2% | 65.5% | 16.0% | 30.8% |
| 5425 | NSP3 | G | T | 0.0% | 1.4% | 29.8% | 10.7% | 25.7% | 7.0% | 29.9% | 0.0% | 27.9% | 19.4% | 0.0% | 20.2% |
| 8389 | NSP3 | C | T | 96.4% | 0.2% | 65.6% | 5.2% | 53.6% | 79.5% | 65.5% | 97.8% | 64.4% | 60.8% | 98.9% | 42.4% |
| 9130 | NSP4 | G | T | 0.0% | 16.9% | 1.3% | 21.9% | 0.0% | 0.3% | 0.6% | 0.0% | 1.4% | 1.1% | 0.0% | 0.7% |
| 10097 | NSP5 | A | G | 15.8% | 0.1% | 0.0% | 0.3% | 19.7% | 10.9% | 0.0% | 16.0% | 0.0% | 0.3% | 0.0% | 18.9% |
| 10700 | NSP5 | G | A | 1.7% | 63.8% | 87.6% | 73.1% | 16.7% | 67.0% | 17.9% | 87.5% | 25.6% | 9.7% | 0.0% | 58.9% |
| 11770 | NSP6 | A | G | 12.2% | 96.9% | 4.4% | 79.1% | 0.1% | 0.8% | 3.3% | 13.6% | 5.6% | 4.8% | 0.1% | 0.7% |
| 12043 | NSP7 | C | T | 44.1% | 93.2% | 5.2% | 78.4% | 0.5% | 0.2% | 3.3% | 44.2% | 5.5% | 4.4% | 0.2% | 0.4% |
| 13527 | NSP12b | T | C | 0.0% | 2.6% | 27.6% | 15.1% | 13.0% | 12.1% | 26.0% | 0.9% | 27.6% | 27.9% | 18.7% | 12.9% |
| 14776 | NSP12b | G | T | 1.4% | 50.2% | 1.5% | 32.6% | 0.0% | 0.2% | 1.0% | 1.3% | 3.3% | 2.3% | 0.0% | 0.0% |
| 16733 | NSP13 | C | T | 0.0% | 51.4% | 2.3% | 31.3% | 0.1% | 0.2% | 1.5% | 0.0% | 1.8% | 0.9% | 0.0% | 0.0% |
| 17320 | NSP13 | G | T | 0.0% | 0.1% | 0.0% | 0.2% | 0.0% | 0.4% | 0.0% | 0.0% | 0.0% | 0.0% | 0.0% | 0.0% |
| 17703 | NSP13 | C | T | 0.0% | 0.1% | 0.2% | 0.1% | 19.0% | 1.1% | 0.0% | 0.1% | 0.1% | 0.2% | 14.2% | 18.9% |
| 18488 | NSP14a | T | C | 0.0% | 9.7% | 0.7% | 17.1% | 0.0% | 1.2% | 1.0% | 0.0% | 0.8% | 2.0% | 0.0% | 1.7% |
| 19388 | NSP14a | G | A | 0.0% | 7.6% | 1.0% | 16.3% | 0.0% | 0.4% | 0.2% | 0.0% | 0.6% | 1.3% | 0.0% | 0.2% |
| 20150 | NSP15 | A | G | 0.0% | 9.8% | 0.9% | 20.0% | 0.0% | 1.0% | 1.0% | 0.0% | 1.6% | 1.4% | 0.0% | 0.0% |
| 21600 | S | G | T | 0.0% | 2.0% | 31.6% | 16.8% | 45.1% | 17.1% | 35.5% | 0.0% | 28.6% | 31.3% | 2.1% | 40.9% |
| 21635 | S | C | A | 12.1% | 96.5% | 16.9% | 78.6% | 0.0% | 0.4% | 18.9% | 20.0% | 6.1% | 0.0% | 0.0% | 0.0% |
| 21753 | S | T | G | 0.0% | 20.2% | 5.3% | 23.4% | 0.0% | 0.7% | 12.2% | 7.7% | 0.0% | 0.0% | 0.0% | 0.0% |
| 21768 | S | | - | 0.0% | 3.6% | 0.0% | 0.0% | 0.0% | 3.1% | 0.0% | 0.0% | 0.0% | 0.7% | 0.0% | 0.0% |
| 22088 | S | C | T | 67.7% | 90.0% | 0.0% | 0.0% | 70.3% | 96.5% | 0.0% | 0.0% | 86.3% | 91.0% | 0.0% | 0.0% |
| 22160 | S | T | C | 0.0% | 3.0% | 0.0% | 5.9% | 0.0% | 1.4% | 0.0% | 0.0% | 0.2% | 0.0% | 0.0% | 6.3% |
| 22281 | S | C | T | 9.1% | 1.4% | 0.0% | 13.9% | 23.9% | 8.1% | 35.7% | 0.0% | 28.2% | 16.7% | 0.8% | 33.3% |
| 22551 | S | C | T | 5.7% | 1.0% | 0.0% | 10.7% | 26.7% | 8.0% | 6.1% | 0.0% | 26.6% | 3.0% | 0.8% | 35.0% |

| | | | | | | | | | | | | | | | |
|---|---|---|---|---|---|---|---|---|---|---|---|---|---|---|---|
| 23398 | S | T | C | 20.0% | 0.0% | 0.0% | 0.0% | 0.0% | 4.8% | 0.0% | 0.0% | 0.0% | 0.0% | 0.0% | 0.0% |
| 23948 | S | G | C | 0.0% | 1.8% | 36.9% | 13.0% | 35.1% | 10.0% | 30.5% | 0.0% | 26.6% | 28.5% | 0.1% | 28.1% |
| 24257 | S | G | T | 82.7% | 0.0% | 58.0% | 6.8% | 72.2% | 81.2% | 71.8% | 85.6% | 66.5% | 70.8% | 100.0% | 76.7% |
| 25537 | E | G | A | 0.0% | 0.7% | 0.0% | 0.2% | 10.1% | 0.3% | 0.0% | 0.0% | 0.1% | 0.2% | 0.0% | 7.4% |
| 26333 | E | C | T | 0.0% | 0.3% | 0.0% | 0.1% | 0.0% | 0.3% | 0.0% | 0.1% | 0.0% | 0.1% | 0.0% | 0.0% |
| 26634 | M | G | T | 38.6% | 96.3% | 0.9% | 76.9% | 0.0% | 0.1% | 3.7% | 40.9% | 7.9% | 7.7% | 0.0% | 1.0% |
| 26647 | M | G | A | 1.8% | 30.9% | 0.2% | 9.2% | 0.0% | 0.4% | 0.0% | 2.0% | 0.6% | 0.4% | 0.0% | 0.0% |
| 27087 | M | G | A | 0.0% | 25.4% | 0.2% | 5.3% | 0.0% | 0.1% | 0.1% | 0.1% | 0.0% | 0.4% | 0.1% | 0.0% |
| 27408 | ORF7a | T | C | 10.0% | 10.9% | 1.0% | 19.4% | 0.0% | 0.2% | 0.8% | 7.6% | 1.2% | 0.9% | 0.0% | 0.0% |
| 27459 | ORF7a | G | T | 6.4% | 2.0% | 29.7% | 10.3% | 16.9% | 7.1% | 32.3% | 3.6% | 26.9% | 22.5% | 0.0% | 14.0% |
| 27509 | ORF7a | C | T | 76.5% | 0.4% | 64.1% | 6.9% | 50.5% | 74.7% | 59.6% | 80.9% | 54.6% | 65.1% | 79.7% | 55.9% |
| 27618 | ORF7a | T | C | 0.1% | 0.3% | 0.0% | 0.5% | 1.2% | 0.5% | 0.1% | 0.3% | 0.8% | 0.3% | 0.0% | 1.9% |
| 28209 | ORF7b | G | C | 33.3% | 96.4% | 34.8% | 82.8% | 0.0% | 0.0% | 7.2% | 0.0% | 13.0% | 0.0% | 0.0% | 0.0% |
| 28356 | N | A | G | 0.0% | 1.8% | 25.7% | 13.2% | 1.3% | 2.6% | 28.5% | 0.1% | 27.5% | 25.0% | 0.0% | 0.8% |
| 28748 | N | C | T | 87.9% | 0.6% | 55.1% | 7.2% | 50.0% | 78.6% | 58.9% | 86.5% | 56.4% | 59.0% | 94.6% | 45.0% |
| 29825 | 3'UTR | G | T | 0.0% | 2.4% | 26.1% | 13.5% | 0.1% | 3.3% | 27.7% | 0.0% | 26.4% | 30.6% | 0.0% | 0.5% |

**Supplementary Table 5: Single genome sequencing (SGS) data from respiratory samples at indicated days.** Indicated are the number of single genomes obtained at each time point with the mutations of interest (identified by deep sequencing). *denominator is 19 as for 2 samples the primer reads were poor quality at amino acid 796 at day 98.

| | W64G | P330S | 69/70 | D796H | T200I | Y240H |
|---|---|---|---|---|---|---|
| Day 1 (n=7) | 0 | 0 | 0 | 0 | 0 | 0 |
| Day 37 (n=38) | 0 | 0 | 0 | 0 | 0 | 0 |
| Day 98 (n=21) | 1 (4.8%) | 1 (4.8%) | 17 (81.0%) | 13*(68.4%) | 3 (14.3%) | 3 (14.3%) |

**Supplementary table 6: Neutralisation of mutants by *Seven RBD-specific mAbs (from Bauwer et al. in Figure 7).*** Clusters II, V contain only non-neutralising mAbs, smaller neutralising mAb clusters IV (n=2) and X (n=1) were not tested. Red indicates significant fold changes.

| mAb | Cluster | Target | | WT | D796H | Δ6970 | Δ6970-D796H | Fold decrease | | |
|---|---|---|---|---|---|---|---|---|---|---|
| COVA1-18 | I | RBD | | 0.0014 | 0.0022 | 0.0016 | 0.0013 | 1.6 | 1.2 | 1.0 |
| COVA2-39 | I | RBD | Structure | 0.0143 | 0.0203 | 0.0319 | 0.0163 | 1.4 | 2.2 | 1.1 |
| COVA1-16 | III | RBD | Structure | 0.2441 | 0.1242 | 0.2651 | 0.1308 | 0.5 | 1.1 | 0.5 |
| COVA2-07 | III | RBD | | 0.0349 | 0.0269 | 0.0288 | 0.0272 | 0.8 | 0.8 | 0.8 |
| COVA2-04 | III | RBD | Structure | 0.2887 | 0.1009 | 0.2425 | 0.1401 | 0.3 | 0.8 | 0.5 |
| COVA2-17 | IX | RBD | | 0.0156 | 0.0248 | 0.0139 | 0.0113 | 1.6 | 0.9 | 0.7 |
| COVA2-02 | VII | RBD | | 5.8590 | 4.9670 | 6.5680 | 3.5380 | 0.8 | 1.1 | 0.6 |
| COVA1-12 | VI | RBD | | 0.2007 | 0.1863 | 0.1105 | 0.0611 | 0.9 | 0.6 | 0.3 |
| COVA1-21 | XI | Non-RBD | | 0.1189 | 0.0498 | 0.6035 | 0.5682 | 0.4 | 5.1 | 4.8 |

**Supplementary Table 7. Global prevalence of selected spike mutations detailed in this paper.** All high coverage sequences were downloaded from the GISAID database on 11th November and aligned using MAFFT. The global prevalence of each of the six spike mutations W64G, ΔH69/V70, Y200H, T240I, P330S and D796H were assessed by viewing the multiple sequence alignment in AliView, sorting by the column of interest, and counting the number of mutations.

| Mutation | Number of Sequences | Global Prevalence |
|---|---|---|
| W64G | 0 | 0.00% |
| ΔH69/V70 | 2920 | 1.38% |
| Y200H | 7 | 0.00% |
| T240I | 34 | 0.02% |
| P330S | 130 | 0.06% |
| D796H | 25 | 0.01% |

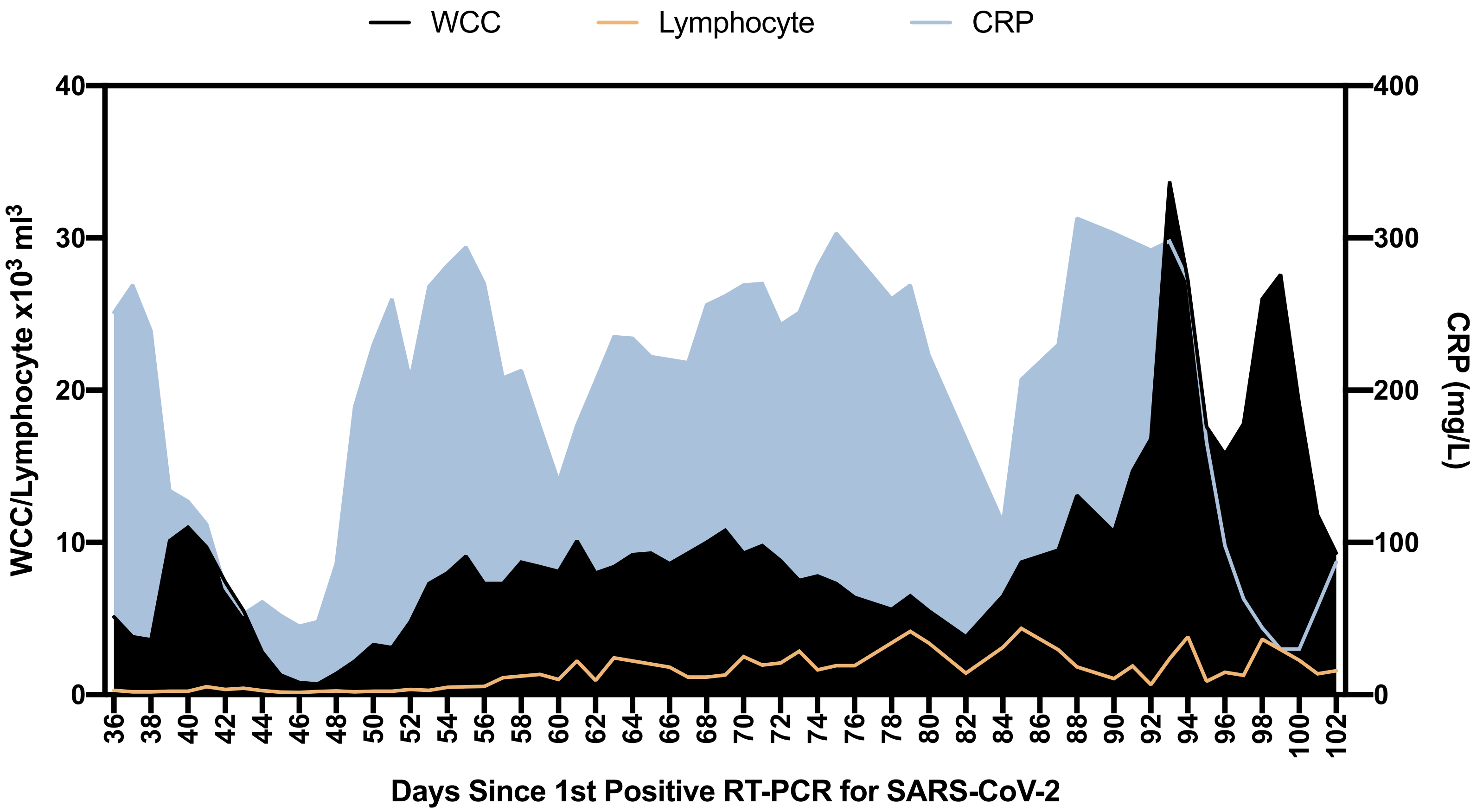


**Supplementary Figure 1: Blood parameters over time in patient case:** White cell count (WCC) and lymphocyte counts are expressed as x$10^3$ Cells/$mm^3$. CRP: C reactive protein

**Supplementary Figure 2: Assessment of T cell and innate function.** Whole blood cytokines were measured in whole blood after 24 hours stimulation either after T-cell stimulation with PHA or anti CD3/IL2 or innate stimulation with LPS . Healthy controls are shown as grey circles (N=15), Patient at d71 and d98 is shown as blue circles or red circles respectively. Cytokine levels are shown as pg/ml. stimulation.

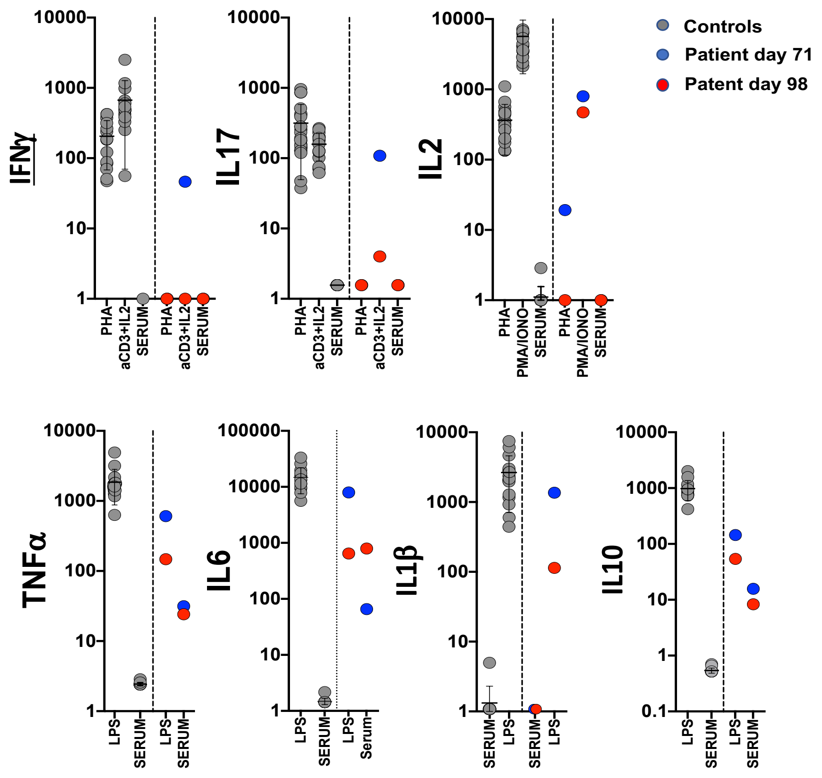

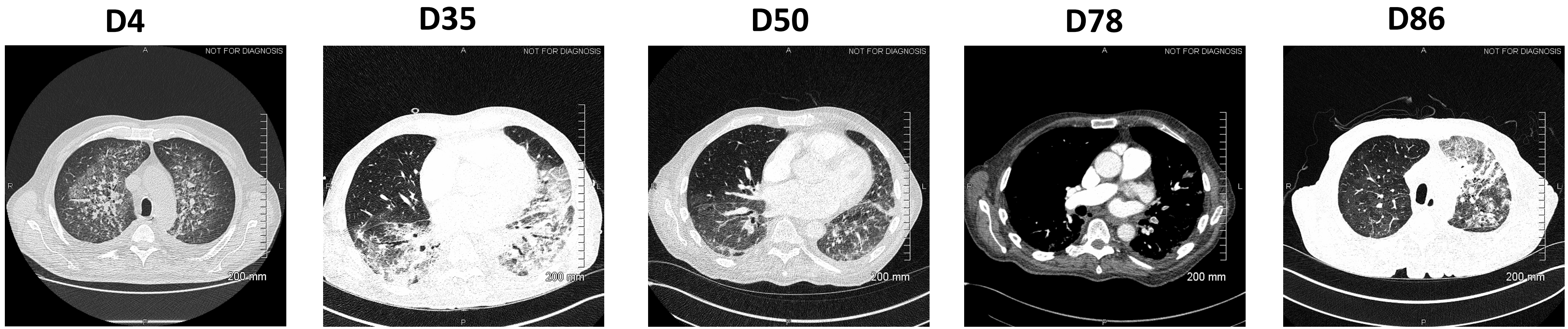


**Supplementary Figure 3: Serial CT images following detection of SARS-CoV-2.**

The patient initially presented with ground glass and peribroncho-vascular consolidation with associated intralobular septal thickening/reticulation and architectural distortion and interlobular septal thickening. By day 50 there is some improvement with evidence of resolving pnuemonitis, however, his condition deteriorated following the detection of bilateral pulmonary emboli, a well-recognized complication of SARS-CoV-2. Despite multiple therapeutic interventions, the patient’s condition deteriorated with worsening of inflammatory changes and chronic organizing pneumonia (COP), particularly on the left, and ongoing changes compatible with persistent SARS-CoV2 infection.

**Supplementary Figure 4: SARS-CoV-2 antibody titres in convalescent plasma.** Measurement of SARS-CoV-2 specific IgG antibody titres in three units of convalescent plasma (CP) by Euroimmun assay.

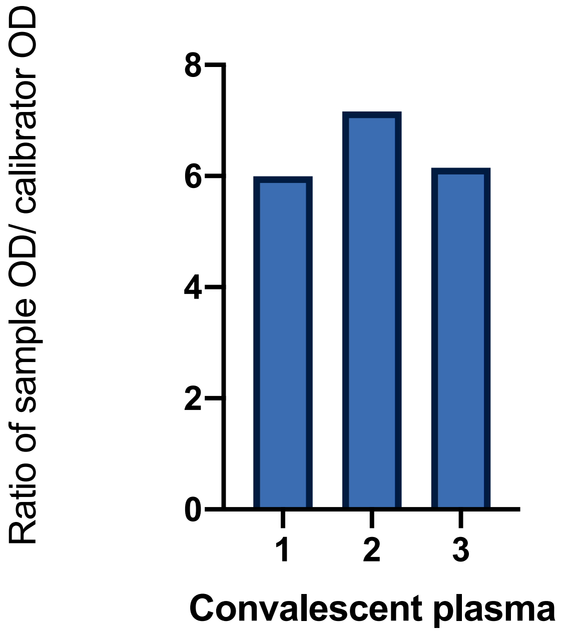

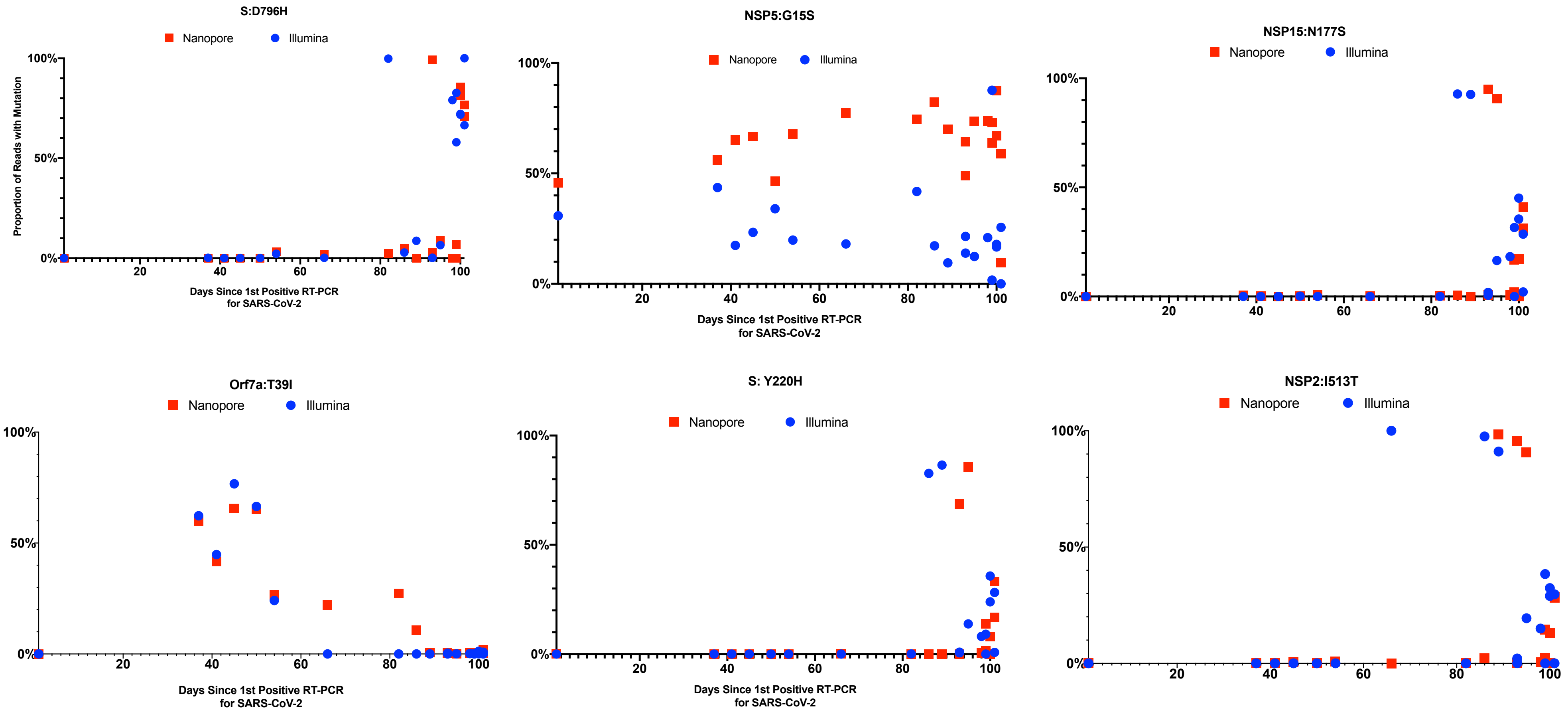


**Supplementary Figure 5. Concordance between short-read (Illumina) and long-read single molecule (Oxford Nanopore) sequencing methods for twenty samples.** Points are the frequency of the variant at each timepoint; only 20 samples were sequenced using the Illumina method. Boxes represent inter-quartile ranges. Error bars are 95% CI. There was good concordance for mutations between the two methods, and no significant difference between the proportion of reads measured by both methods.

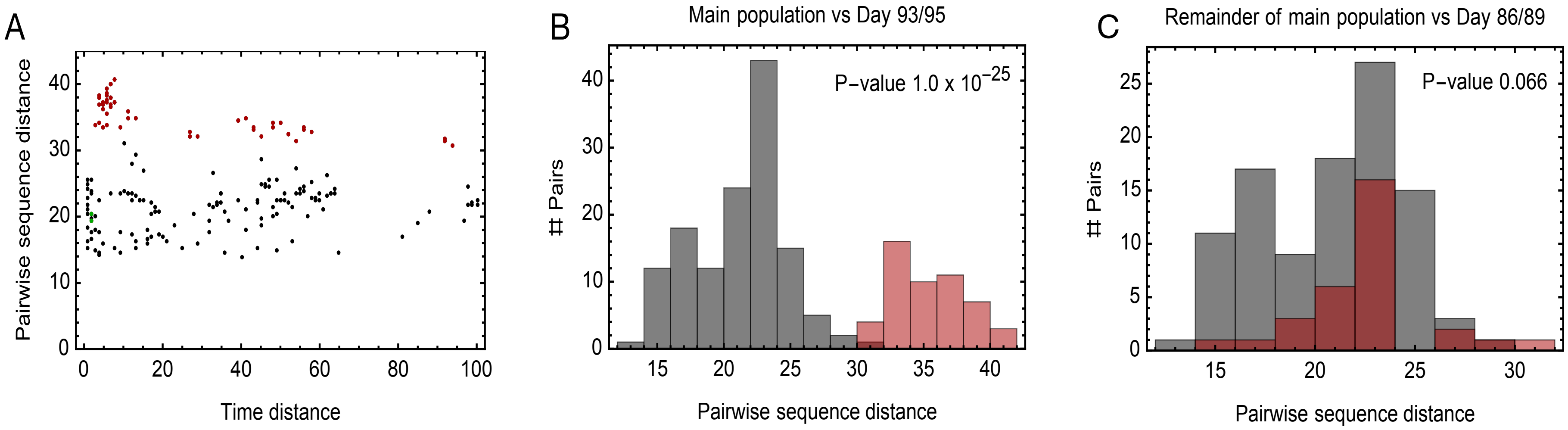


**Supplementary Figure 6: Additional evidence for within-host cladal structure. A.** Pairwise distances between samples measured using the all-locus distance metric plotted against pairwise distances in time (measured in days) between samples being collected. Internal distances between samples in the proposed main clade are shown in black, distances between samples in the main clade and samples collected on days 93 and 95 are shown in red, and internal distances between samples collected on days 93 and 95 are shown in green. **B.** Pairwise distances between samples in the larger clade (black) and between these samples and those collected on days 93 and 95 (red). The median values of the distributions of these values are significantly different according to a Mann Whitney test. **C.** Pairwise distances between samples in the main clade, once those collected on days 86, 89, 93, 95 have been removed (black) and between these samples and those collected on days 86 and 89 (red). The median values of the distributions of these values are not significantly different at the 5% level according to a Mann Whitney test.

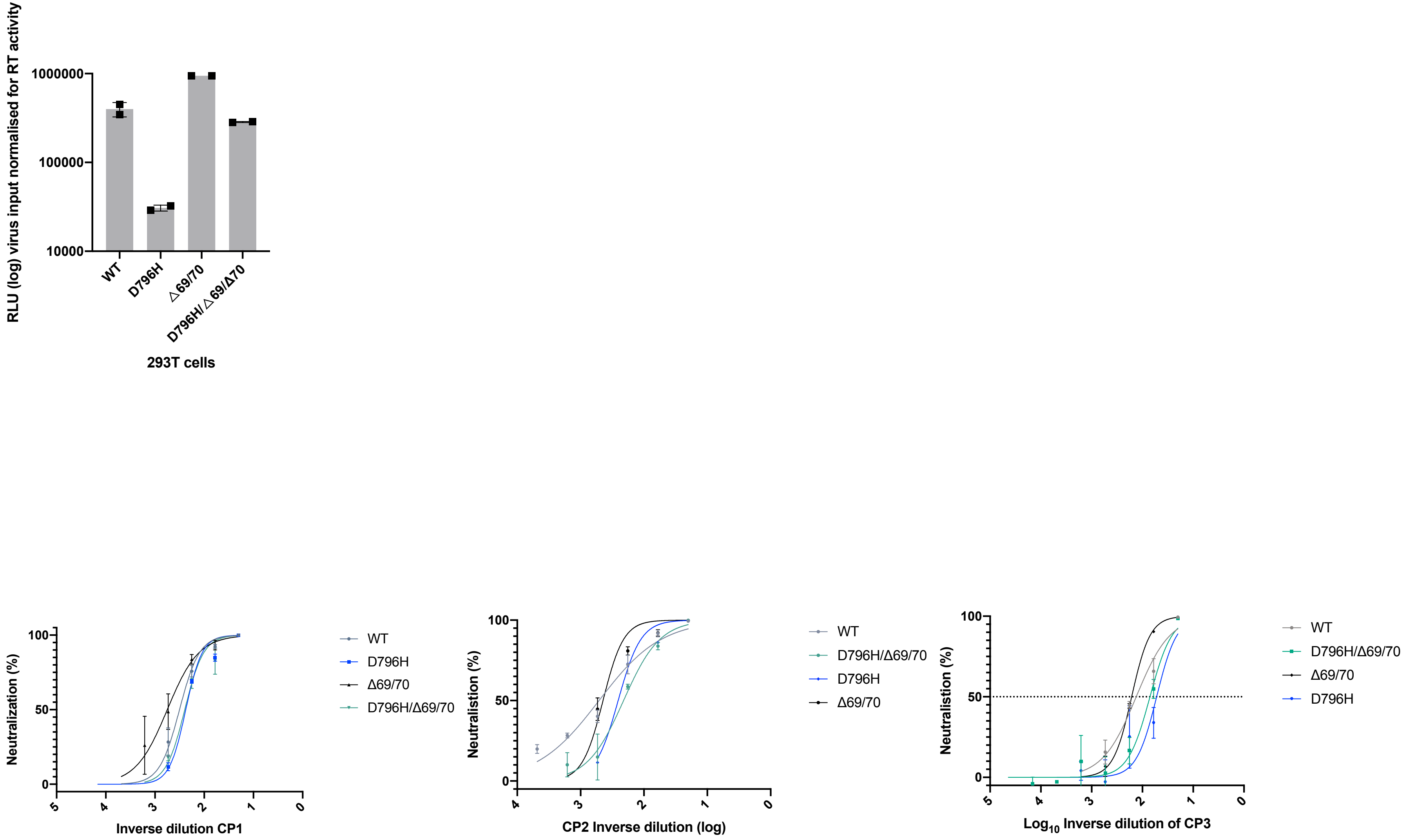


**Supplementary Figure 7: In vitro infectivity and neutralisation sensitivity of Spike pseudotyped lentiviruses.** Top: infection of target 293T cells expressing TMPRSS2 and ACE2 receptors using equal amounts of virus as determined by reverse transcriptase activity. Bottom: Representative Inverse dilution plots for Spike variants against convalescent plasma units 1-3. Data points represent mean % neutralisation and error bars represent standard error of the mean
